# Forecasting the impact of the first wave of the COVID-19 pandemic on hospital demand and deaths for the USA and European Economic Area countries

**DOI:** 10.1101/2020.04.21.20074732

**Authors:** IHME COVID-19 health service utilization forecasting team, Christopher JL Murray

## Abstract

**Background:** Hospitals need to plan for the surge in demand in each state or region in the United States and the European Economic Area (EEA) due to the COVID-19 pandemic. Planners need forecasts of the most likely trajectory in the coming weeks and will want to plan for the higher values in the range of those forecasts. To date, forecasts of what is most likely to occur in the weeks ahead are not available for states in the USA or for all countries in the EEA.

**Methods:** This study used data on confirmed COVID-19 deaths by day from local and national government websites and WHO. Data on hospital capacity and utilisation and observed COVID-19 utilisation data from select locations were obtained from publicly available sources and direct contributions of data from select local governments. We develop a mixed effects non-linear regression framework to estimate the trajectory of the cumulative and daily death rate as a function of the implementation of social distancing measures, supported by additional evidence from mobile phone data. An extended mixture model was used in data rich settings to capture asymmetric daily death patterns. Health service needs were forecast using a micro-simulation model that estimates hospital admissions, ICU admissions, length of stay, and ventilator need using available data on clinical practices in COVID-19 patients. We assume that those jurisdictions that have not implemented school closures, non-essential business closures, and stay at home orders will do so within twenty-one days.

**Findings:** Compared to licensed capacity and average annual occupancy rates, excess demand in the USA from COVID-19 at the estimated peak of the epidemic (the end of the second week of April) is predicted to be 9,079 (95% UI 253–61,937) total beds and 9,356 (3,526–29,714) ICU beds. At the peak of the epidemic, ventilator use is predicted to be 16,545 (8,083–41,991). The corresponding numbers for EEA countries are 120,080 (119,183–121,107), 32,291 (32,157– 32,425) and 28,973 (28,868–29,085) at a peak of April 6. The date of peak daily deaths varies from March 30 through May 12 by state in the USA and March 27 through May 4 by country in the EEA. We estimate that through the end of July, there will be 60,308 (34,063–140,381) deaths from COVID-19 in the USA and 143,088 (101,131–253,163) deaths in the EEA. Deaths from COVID-19 are estimated to drop below 0.3 per million between May 4 and June 29 by state in the USA and between May 4 and July 13 by country in the EEA. Timing of the peak need for hospital resource requirements varies considerably across states in the USA and across regions of Europe.

**Interpretation:** In addition to a large number of deaths from COVID-19, the epidemic will place a load on health system resources well beyond the current capacity of hospitals in the USA and EEA to manage, especially for ICU care and ventilator use. These estimates can help inform the development and implementation of strategies to mitigate this gap, including reducing non-COVID-19 demand for services and temporarily increasing system capacity. The estimated excess demand on hospital systems is predicated on the enactment of social distancing measures within three weeks in all locations that have not done so already and maintenance of these measures throughout the epidemic, emphasising the importance of implementing, enforcing, and maintaining these measures to mitigate hospital system overload and prevent deaths.

**Funding:** Bill & Melinda Gates Foundation and the state of Washington

## Introduction

The Coronavirus Disease 2019 (COVID-19) pandemic started in Wuhan, China, in December 2019^1^ and has since spread to the vast majority of countries.^2^ As of April 16, twelve countries have recorded more than a thousand deaths: Italy, USA, Spain, France, UK, Iran, China, Netherlands, Germany, Belgium, Canada, and Switzerland. COVID-19 is not only causing mortality but is also putting considerable stress on health systems, with large case numbers and many patients needing critical care including mechanical ventilation. Estimates of the potential magnitude of COVID-19 patient volume – particularly at the local peak of the epidemic –are urgently needed for USA and European hospitals still early in the epidemic to effectively manage the rising case load and provide the highest quality of care possible.

COVID-19 scenarios and forecasts have largely been based on mathematical compartmental models that capture the probability of moving between susceptible, exposed, and infected states, and then to a recovered state or death (SEIR models). Many SEIR or SIR models have been published or posted online.^3–20^ In general, these models assume random mixing between all individuals in a given population. While results of these models are sensitive to starting assumptions and thus differ between models considerably, they generally suggest that given current estimates of the basic reproductive rate (the number of cases caused by each case in a susceptible population), 25% to 90% of the population could eventually become infected unless mitigation measures are put in place and maintained.^6,20^ Based on reported case-fatality rates, these projections imply that there would be millions of deaths in the USA and Europe due to COVID-19. Individual behavioural responses and government-mandated social distancing (school closures, non-essential service closures, and shelter-in-place orders), however, can dramatically influence the course of the epidemic. As of April 14, 2020, for Wuhan City in China – and also for at least 12 additional regions in Italy (Liguria, Lombardia, Emilia-Romagna, Marche, Lazio, Campania), Spain (Community of Madrid, Castile and Leon, Catalonia, Navarre), and the USA (King County, Snohomish County) – strict social distancing has led to the peak of the first wave of the epidemic, implying that the effective reproduction number (R_effective_) has dropped below unity in these settings. Planning tools based on SEIR models provide high-level information across populations. Few of these planning models have forecasted peaks in deaths or cases and subsequent declines. Using reported case numbers and models based on those for health service planning is also not ideal because of widely varying COVID-19 testing rates and strategies. For example, countries such as Germany, Iceland, and South Korea have undertaken widespread testing, while in the USA and elsewhere, limited test availability has led to largely restricting testing, particularly early in the epidemic, to those with more severe disease or those who are at risk of serious complications.

An alternative strategy is to focus on modelling the empirically observed COVID-19 population death rate curves, which directly reflect both the transmission of the virus and the infection-fatality rates in each specific community. Deaths are likely more accurately reported than cases in settings with limited testing capacity, where tests are usually prioritised for the more severely ill patients. Hospital service need is likely to be highly correlated with deaths, given predictable disease progression probabilities by age for severe cases. In this study, we use statistical modelling to implement this approach and derive state-specific and country-specific forecasts with uncertainty for deaths and for health service resource needs and compare these to available resources in the USA and countries in the European Economic Area (EEA). This model is regularly updated to incorporate new data for the location of interest as well as data from other locations.

## Methods

The modelling approach in this study is divided into four components: (i) identification and processing of COVID-19 data; (ii) statistical model estimation for population death rates as a function of time since the death rate exceeds a threshold in a location; (iii) predicting time to exceed a given population death threshold in locations early in the pandemic; and (iv) modelling health service utilisation as a function of deaths. Additional information on the determination of hospital resource utilisation and capacity is provided in Appendix A; details on curve fitting methods, quantification of uncertainty, and a full specification of the statistical model are available in Appendix B. This study complies with the Guidelines for Accurate and Transparent Health Estimates Reporting (GATHER) statement.^21^

### Data identification and processing

Local government, national government, and WHO websites, and third-party aggregators^22–26^ were used to identify data on confirmed COVID-19 deaths by day of death at the first administrative level (state or province, hereafter “admin 1”). Data on licensed bed and ICU capacity and average annual utilisation by location were obtained from a variety of sources for most countries to estimate baseline capacities; observed COVID-19 utilisation data were obtained for a range of countries and USA states providing information on inpatient and ICU use or were imputed from available resources (Appendix A). Other parameters were sourced from the scientific literature and an analysis of available patient-level data. Age-specific data on the relative population death rate by age are available from China,^28^ Italy,^29^ South Korea,^30^ the USA,^31,32^ Netherlands,^33^ Sweden,^34^ and Germany^23^ and show a strong relationship with age (Figure 1).

**Figure 1.**
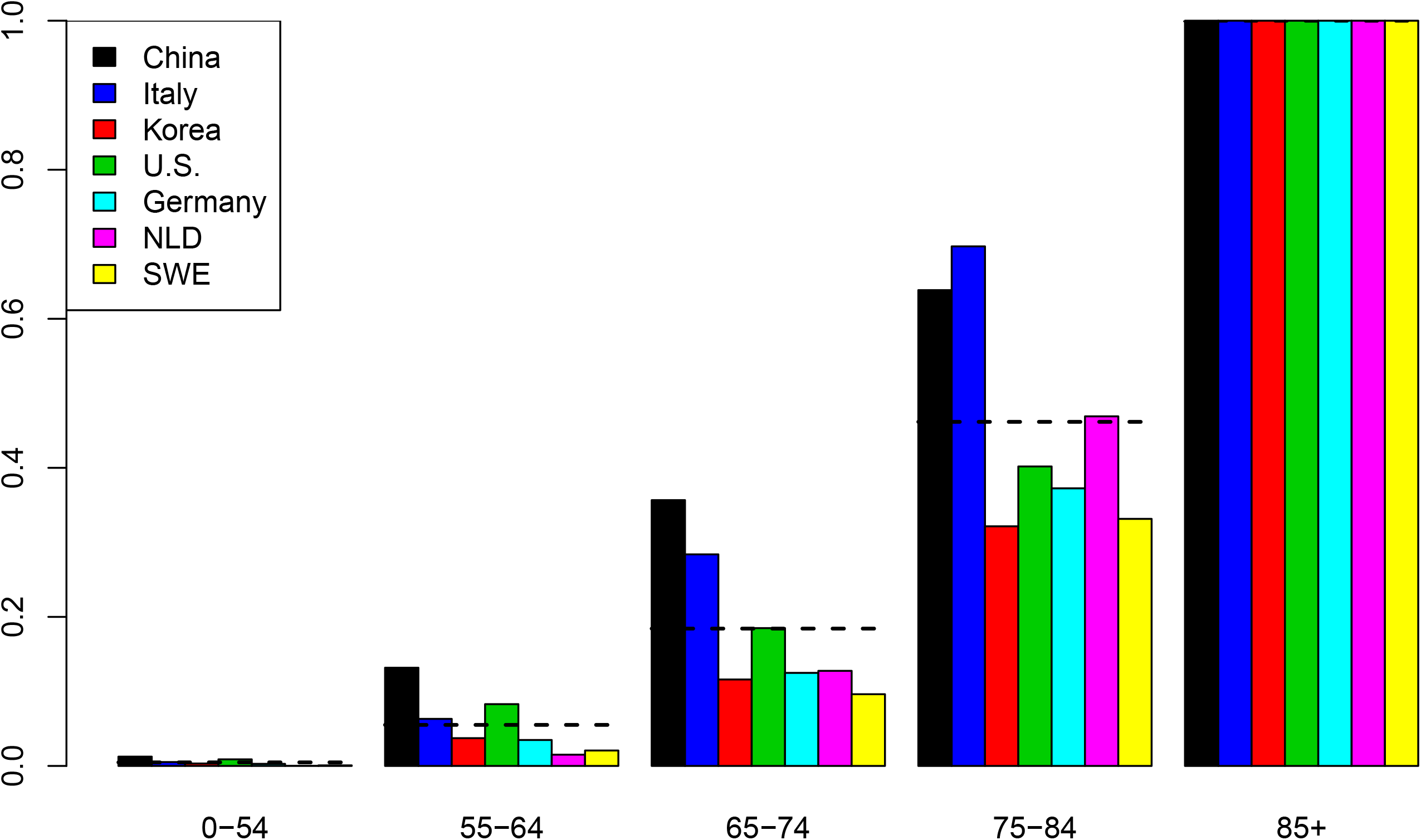
Normalised age-pattern of death based on data from China, Italy, South Korea, Germany, the Netherlands, Sweden, and the USA

Using the average observed relationship between the population death rate and age, data from different locations can be standardised to the age structure using indirect standardisation (Appendix B). For the estimation of statistical models for the population death rate, only admin 1 locations with an observed death rate greater than 0.31 per million (exp(−15)) were used. This threshold was selected by testing which threshold minimised the variance of the slope of the death rate across locations in subsequent days.

Government declarations were used to identify the day that different jurisdictions implemented various social distancing policies (school closures, closures of non-essential services focused on bars and restaurants, stay-at-home or shelter-in-place orders, and the deployment of severe travel restrictions) following the New Zealand government COVID-19 alert schema.^35^ Data on timings of interventions were compiled by checking national and state governmental websites, executive orders, and newly initiated COVID-19 laws, and cross-referencing other policy compilation resources (see Supplementary Information). Covariates of days with expected exponential growth in the cumulative death rate were created using information on the number of days after the death rate exceeded 0.31 per million that six different social distancing measures were mandated by local and national governments: school closures, partial non-essential business closures, complete non-essential business closures, restricting group gatherings, stay-at-home recommendations, and severe local travel restrictions including public transport closures. To derive weighting schemes for each of the social distancing mandates, we determined the effect of social distancing measures on mobility data published by Google (average of retail, workplace, and transit mobility dimensions),^36^ Descartes Lab (distance travelled)^37^ and Safegraph (time spent at home)^38^ using random effects regression where the dependent variable was the log of mobility measures with social distancing measures as a series of dummy variables. The three different weighting schemes were used to create covariates for an ensemble of three models (Appendix B, section 5). For locations that have not yet implemented all of the closure measures, we assumed that the remaining measures will be put in place within 3 weeks. This lag between reaching a threshold death rate and implementing more aggressive social distancing was combined with the observed period of exponential growth in multiple locations that reached their peak after Level 4 social distancing from the New Zealand alert schema^35^ was implemented, adjusted for the median time from incidence to death. For ease of interpretation of statistical coefficients, this covariate was normalised so the value for Wuhan was 1.

### Statistical model for the cumulative death rate

We developed a curve-fitting tool to fit a nonlinear mixed effects model to the available admin 1 cumulative death data. See Appendix B: Curvefit Tool and Analyses for greater detail. The cumulative death rate for each location is assumed to follow a parametrised Gaussian error function:

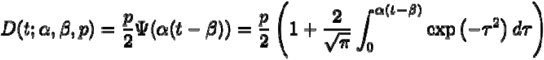

where the function Ψ is the Gaussian error function (written explicitly above), *p* controls the maximum cumulative death rate at each location, *t* is the time since death rate exceeded exp(−15), *ß* (beta) is a location-specific inflection point (time at which rate of increase of the daily death rate is maximum), and α (alpha) is a location-specific growth parameter. Other sigmoidal functional forms (alternatives to Ψ) were considered but did not fit the data as well. Data were fit to the log of the death rate in the available data, using an optimisation framework described in Appendix B. For data-rich cases, we also developed linear curve fitting extension, where after a Gaussian curve in daily death is obtained, we fit the data to a weighted combination (with constraints on weights) of such curves propagated forward and backward in time. The resulting models can capture more complex behavior in the data.

An ensemble of three models was used to produce the estimates. In all models, we parametrised the time-axis shift parameter beta to depend on a covariate based on time from when the initial ln(death rate) exceeds exp(−15) to the implementation of social distancing. The models differed by the definition of the social distancing covariate. In each model, the value of the covariate multiplier was obtained by fitting a joint model on all the locations that were considered to have peaked; that is, the generalisable information from these locations was the impact that social distancing had on the time to reach the inflection point. Using 13 locations where peak deaths had occurred as of April 14, 2020 – China (Wuhan City), Italy (Liguria, Lombardia, Emilia-Romagna, Marche, Lazio, Campania), Spain (Community of Madrid, Castile and Leon, Catalonia, Navarre), and the USA (King County, Snohomish County) – we fit mixed effects models to get the mean and variance of the relationship between the social distancing covariates and the peak time, and used this information to build priors for location-specific estimates.

We use hospitalization data to generate additional short-term predicted deaths (pseudo-data). On average, the time between hospitalization and death is 8 days. Using location-specific hospitalization data which has more than 10 deaths, we estimate the ratio of cumulative deaths to cumulative hospitalizations up to 8 days in the past. We use this ratio to generate pseudo-data for 8 days, and incorporate this pseudo-data into the CurveFit model. Details are given in Section 11 of Appendix B.

For locations with fewer than 18 days, we use the following analysis. For each type of model (based on definition of the covariate), we considered both “short-range” and “long-range” variants, to explain existing data and forecast long-term trends, respectively. In the former case, covariate multipliers could deviate from those estimated using peaked locations, while in the latter, the joint model fit from peaked locations had a larger impact on the final covariate multiplier. The two remaining parameters (not modelled using covariates) were allowed to vary among locations to fit location-specific data. Uncertainty for every model was obtained using the predictive validity framework that analyses errors in predicting out-of-sample observations. Using these methods, we obtain model realisations using draws, for both short- and long-term models across the forecast horizon. We then obtain forecasts that linearly interpolate between short-term and long-term models, with next days closely following short-term models and long-term forecasts following long-term models. Finally, we ensemble these draws across the model types (based on the definition of the social distancing covariate).

For locations with 18 or more days, we first fit a long-term model, borrowing strength from peaked locations and obtaining location-specific representative daily deaths Gaussian curves. We then fit a linear combination of 13 of the inferred Gaussian curves from the long-term model, placed two days apart (12 days back from the inferred peak to 12 days forward of the inferred peak). We then ensemble across draws for different model types. See Appendix B (section 11) for full details.

The dataset age-standardised to the age-structure of California is shown in Figure 2.

**Figure 2.**
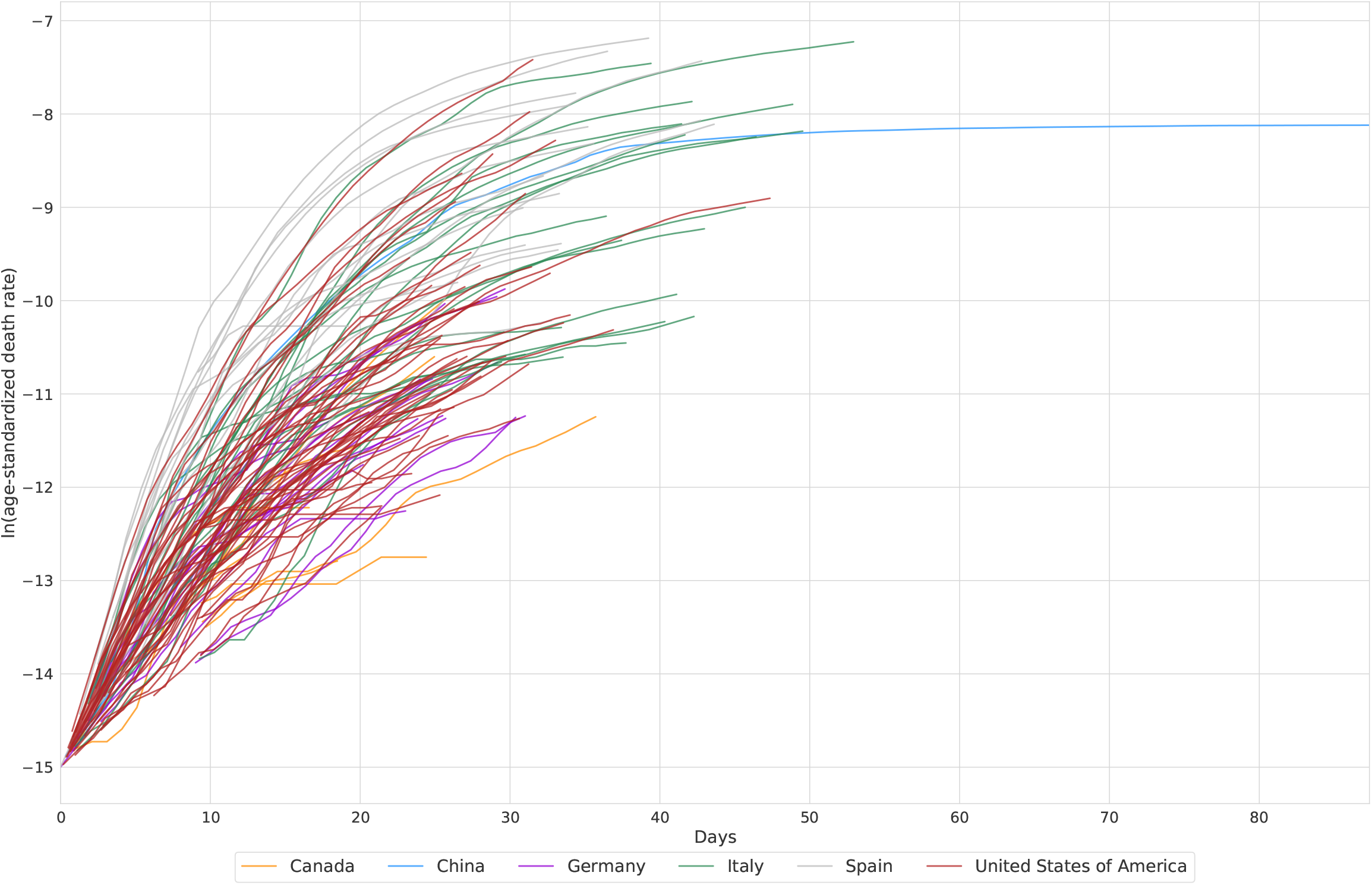
Death rate data age-standardised to California as a function of time since a threshold death rate of 0.3 per million

### Time to threshold death rate

All states except Wyoming have deaths greater than 0.31 per million (e-15) and more than 2 deaths and were included in the model estimation along with data on 66 other admin 1 locations. For other USA states or locations in the EEA, we estimated the expected time from the current case count to reach the threshold level for the population death rate model. Using the observed distribution of the time from each level of case count to the threshold death rate for all admin 1 locations with data, we estimated this distribution. We used the mean and standard deviation of days from a given case count to the future threshold death rate to develop the probability distribution for the day each state will cross over the threshold death rate, and then we applied the death rate epidemic curve after crossing the threshold.

### Hospital service utilisation microsimulation model

From the projected death rates, we estimated hospital service utilisation using an individual-level microsimulation model – additional details are provided in Appendix A. We simulated deaths by age using the average age pattern (Figure 1). For each simulated death, we estimated the date of admission using the median length of stay for deaths from available data (six days). Simulated individuals requiring admission who were discharged alive were generated using the location-specific ratios of admissions to deaths; where location-specific ratios were not available we used the EEA pooled estimate for other EEA countries and the USA pooled estimate for other USA states. An age pattern of the ratio was based on available data (Appendix A). The age-specific fraction of admissions requiring ICU care was based on data from the USA. The fraction of ICU admissions requiring invasive ventilation was estimated as 85%. To determine daily bed and ICU occupancy and ventilator use, we applied median lengths of stay of eight days for those not requiring ICU care and discharged alive and 20 days for those admissions with ICU care, with 13 of those days in the ICU.^39^

## Results

By aggregating forecasts across location, we determined the overall trajectory of expected health-care needs in different categories and deaths, as shown in Figure 3 for the USA (Panel A) and for EEA countries (Panel B). These figures highlight the earlier beginning of the epidemic in EEA countries compared to the USA. The USA projected peak was reached on April 15 with almost 3,500 deaths daily. In EEA the peak was on April 6 with more than 4,000 deaths daily but with a flatter peak, reflecting the considerable variability in the timing of the epidemic by country. Our estimated peak hospital demand was 68,884 (95% UI 34,599–175,312) beds, 18,269 (9,621–44,223) ICU beds and 16,545 (8,083–41,991) ventilators in the USA; for EEA nations the corresponding numbers were 120,080 (119,183–121,107) hospital beds, 32,291 (32,157–32,425) ICU beds, and 28,973 (28,868–29,085) ventilators.

**Figure 3.**
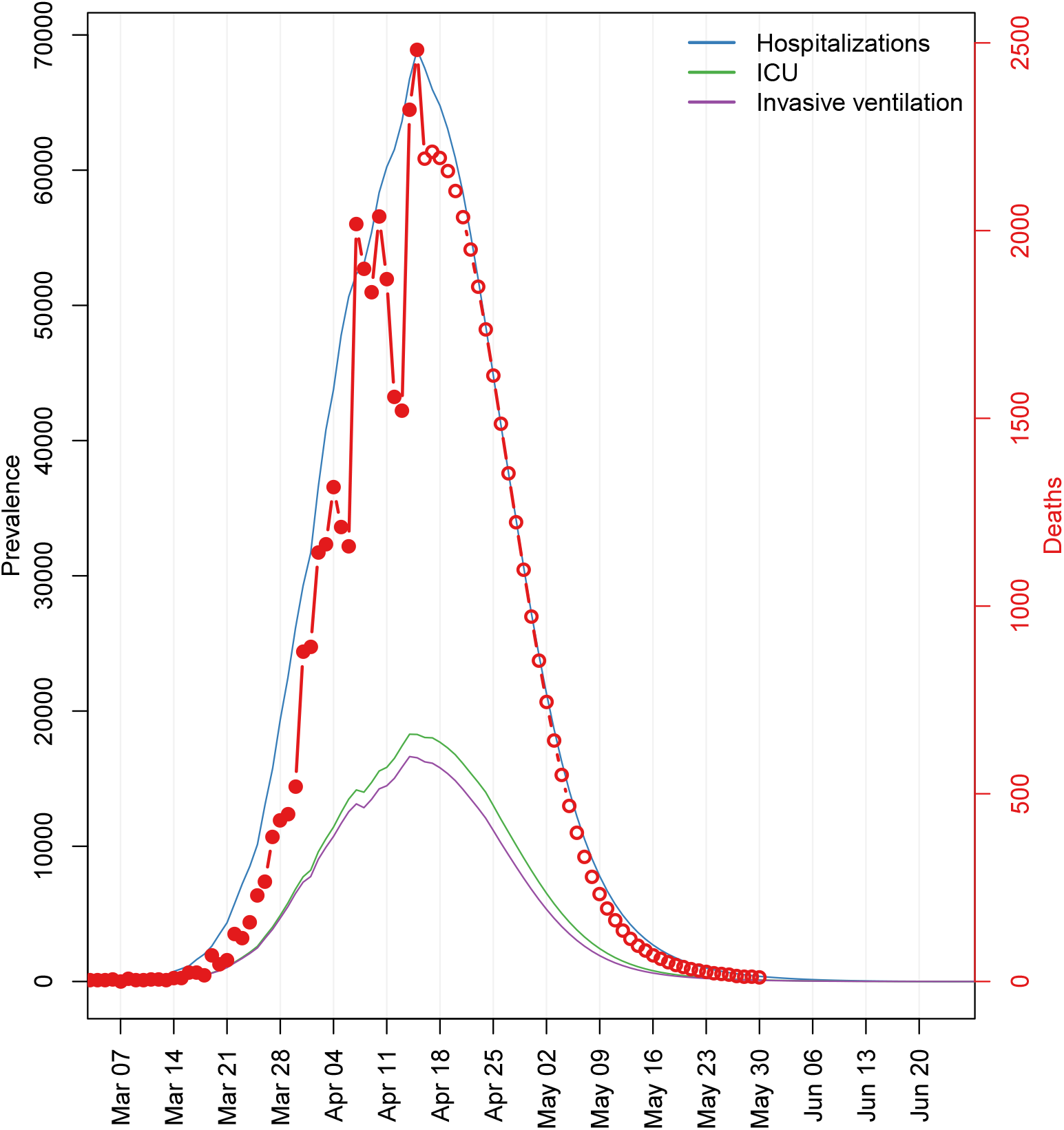
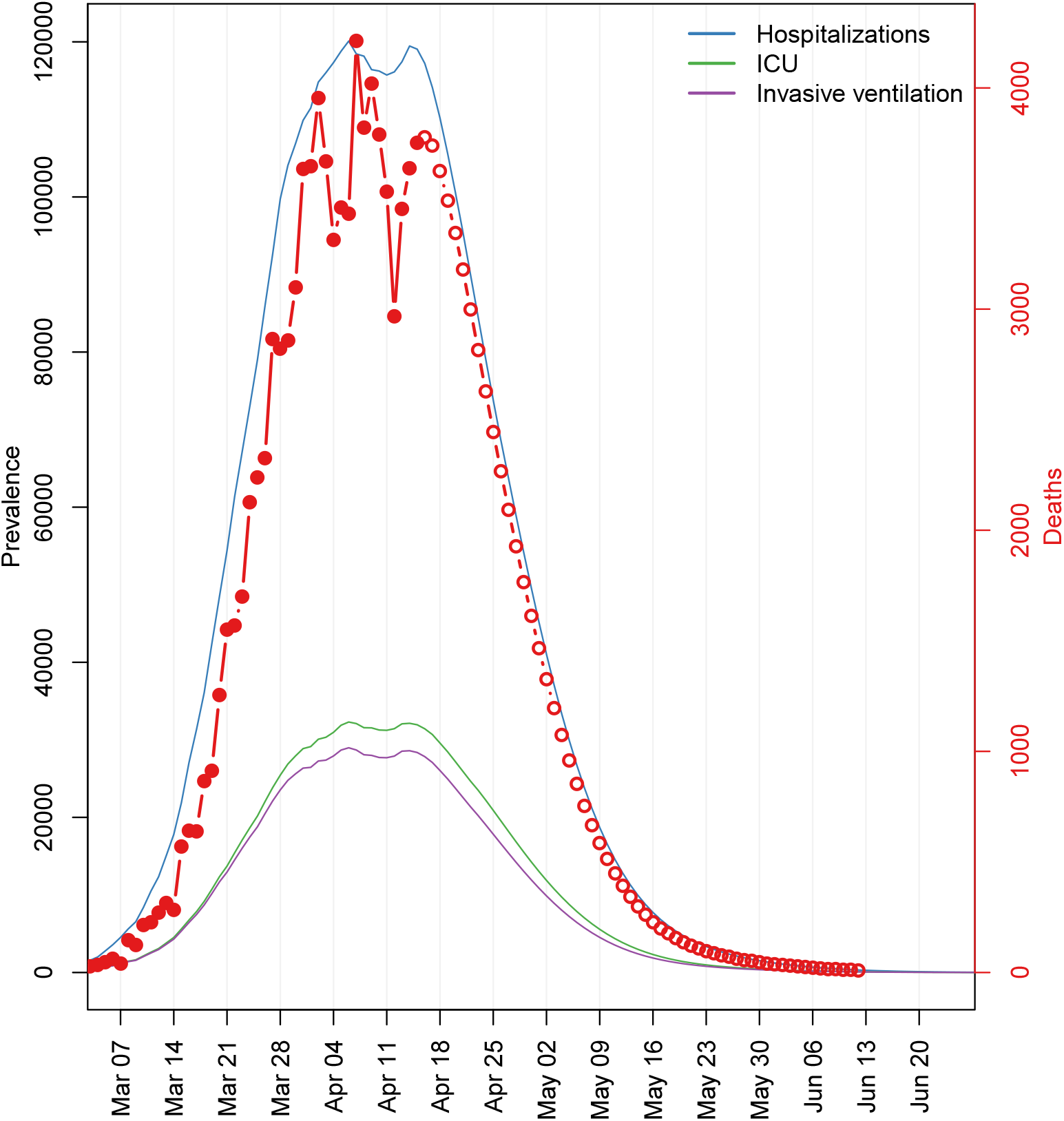
Estimates of hospitalisation and deaths by day for the USA (Panel A) and EEA (Panel B)

The peak of daily deaths varies considerably between EEA countries and subnational locations (Figure 4, Panel A) and USA states (Panel B). Several regions in Italy reached their peak by the end of March, with parts of Spain, France, Netherlands, Norway, Denmark, Greece, and Estonia following suit by the beginning of April. Other countries such as the UK, Germany, and Sweden are at the peak or are approaching the peak. In the USA, states with earlier peaks include Washington, Nevada, Arizona, Montana and Florida. States at the peak or just approaching the peak include Texas, California and parts of New England. States in the middle of the country, including North Dakota, South Dakota, Iowa and Wyoming are expected to peak later.

**Figure 4.**
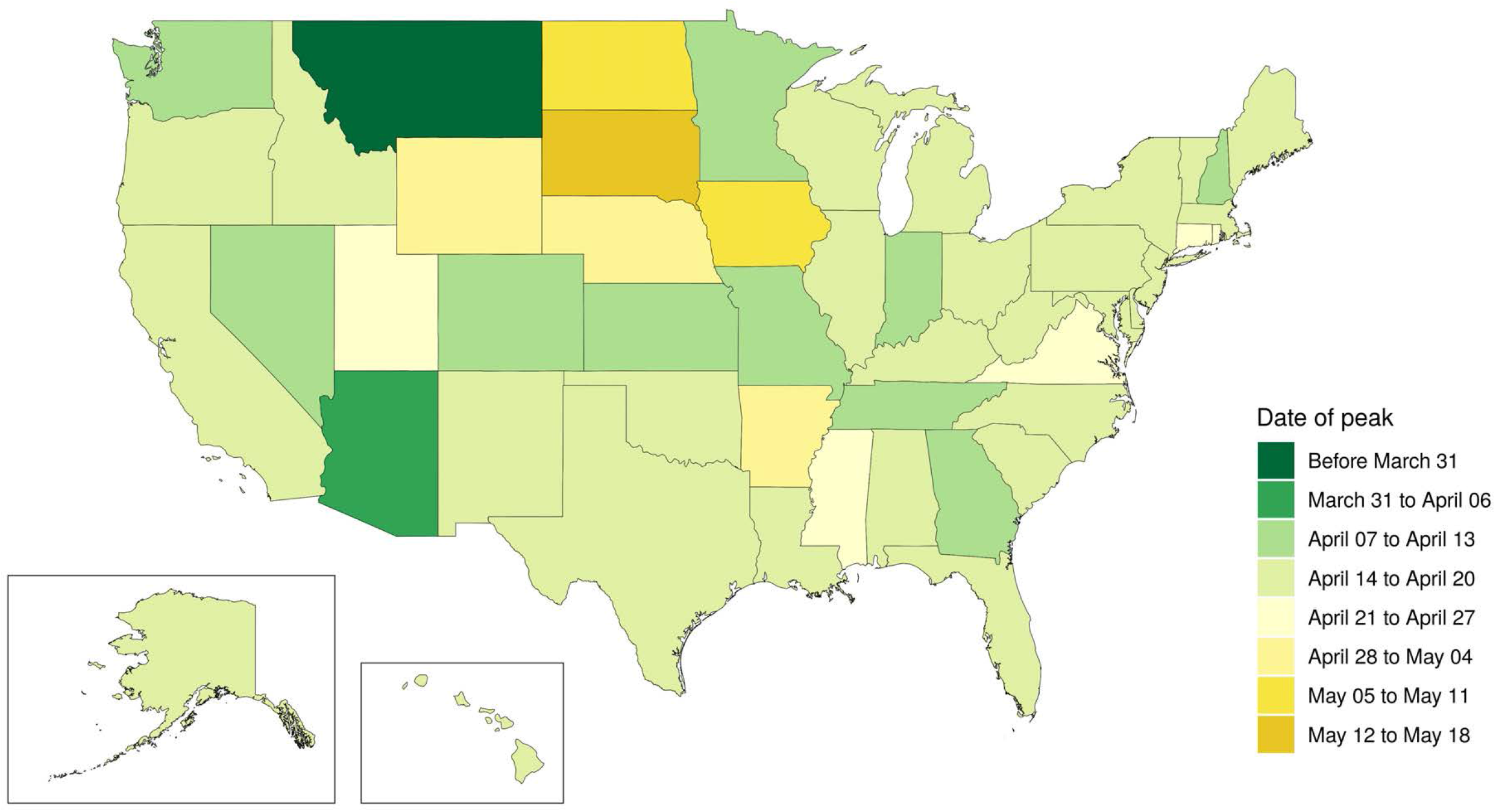
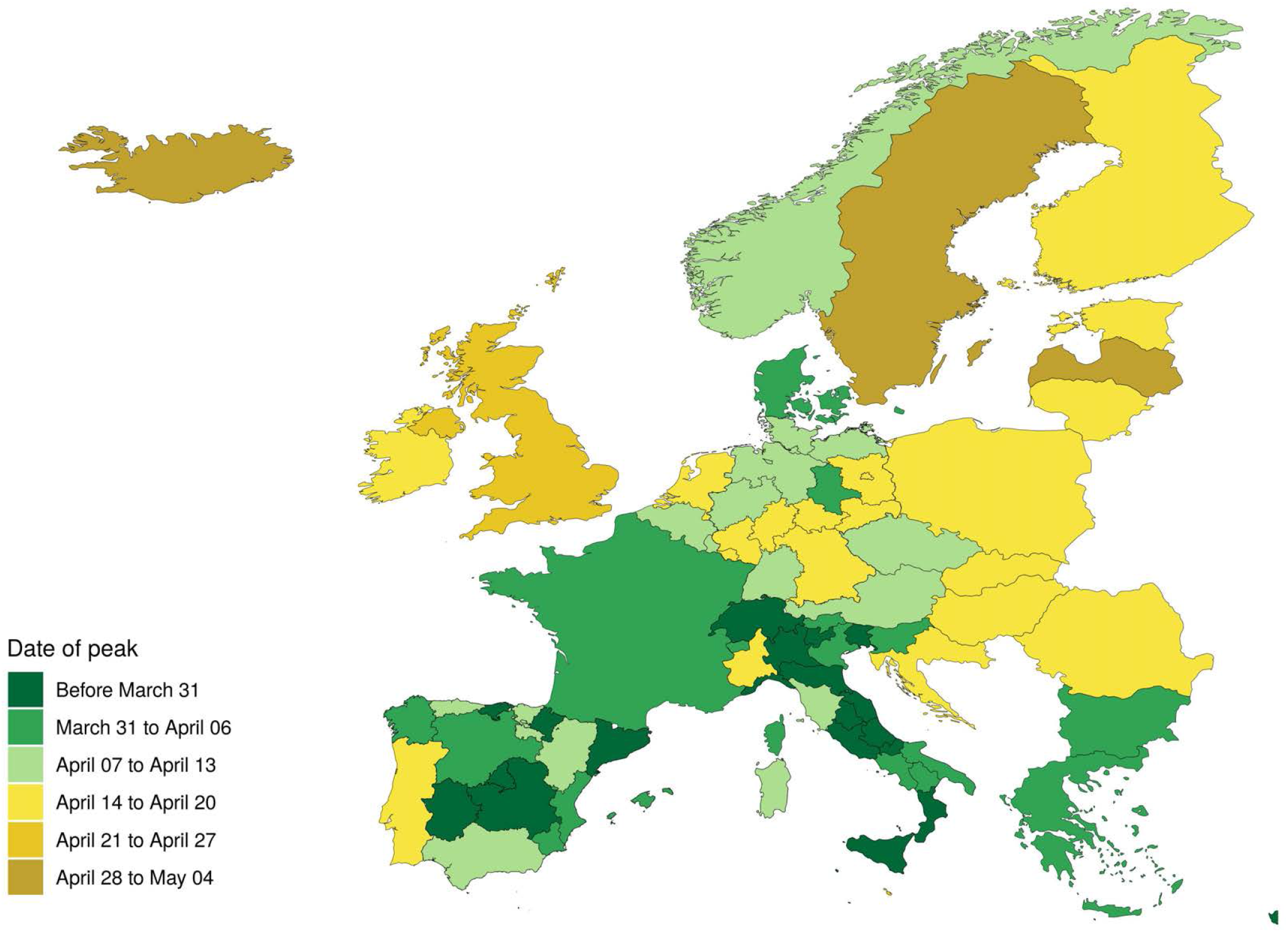
Date of peak daily deaths by location for the USA (Panel A) and EEA (Panel B)

Figure 5 shows total excess demand for the USA (panel A) and EEA countries (panel B) overall. In the USA, peak excess demand for hospitalisation above usual capacity was estimated as 9,079 (95% UI 253–61,937); ICU bed excess demand was 9,356 (3,526–29,714). We estimated that EEA countries experienced a peak excess demand above usual capacity for total beds of 28,270 (0 to 126,788) at peak; the ICU bed shortfall was 16,090 (15,973–16,211). Excess demand is concentrated in particular countries and USA states as shown in Figure 6, which shows the percentage excess demand for ICU beds by location: in the USA (panel A), excess demand for ICU beds is concentrated in New York, New Jersey, Connecticut, Wyoming, Michigan, Rhode Island, and Massachusetts; in the EEA (panel B), ICU excess demand above usual capacity is particularly high in Sweden, Spain, Northern Ireland, Italy, France, and Belgium. We have not been able to estimate current ventilator capacity; however, the number of ventilators per person implied by the peak (Figure 3) also suggests potentially large gaps in availability of ventilators.

**Figure 5.**
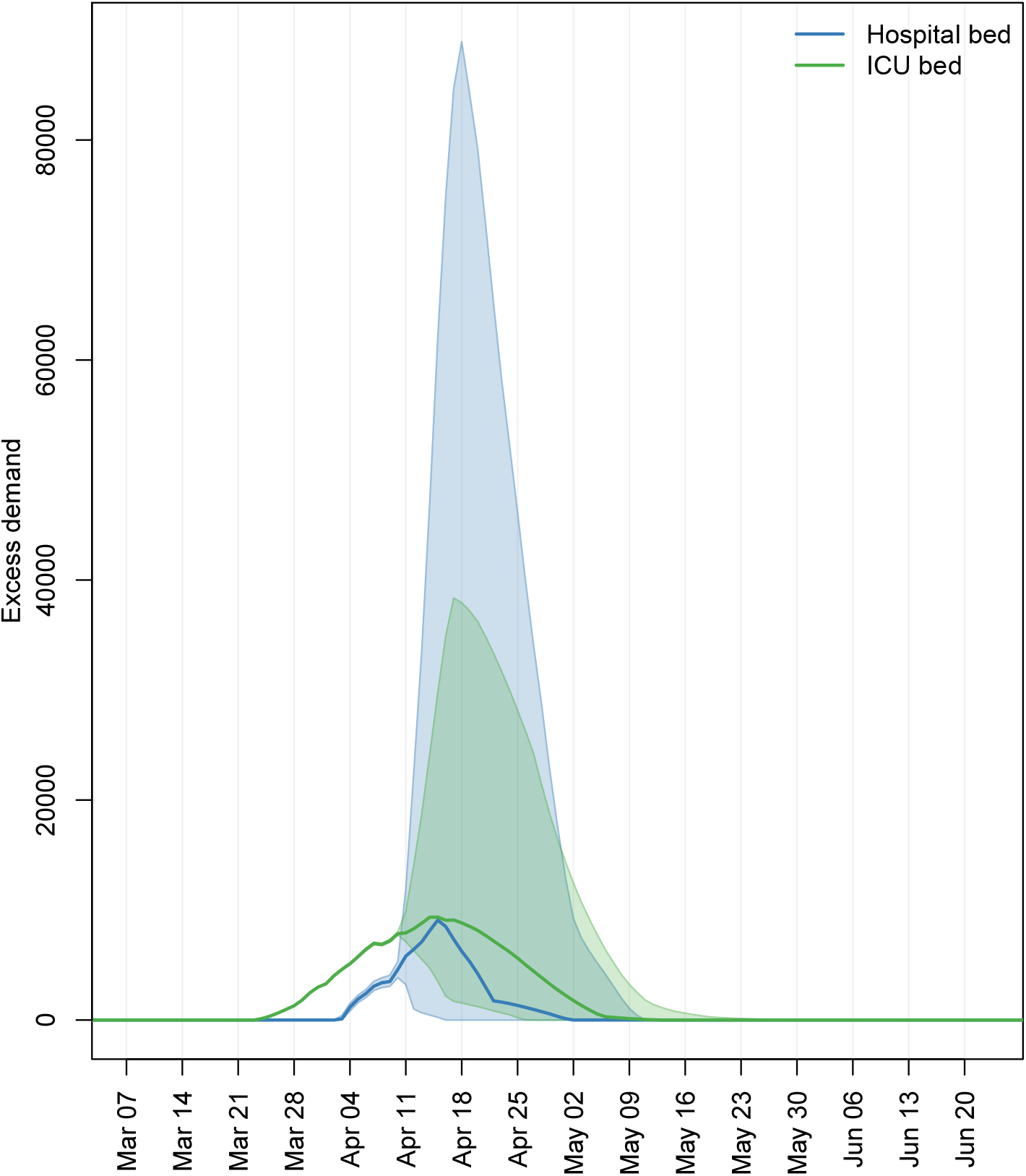
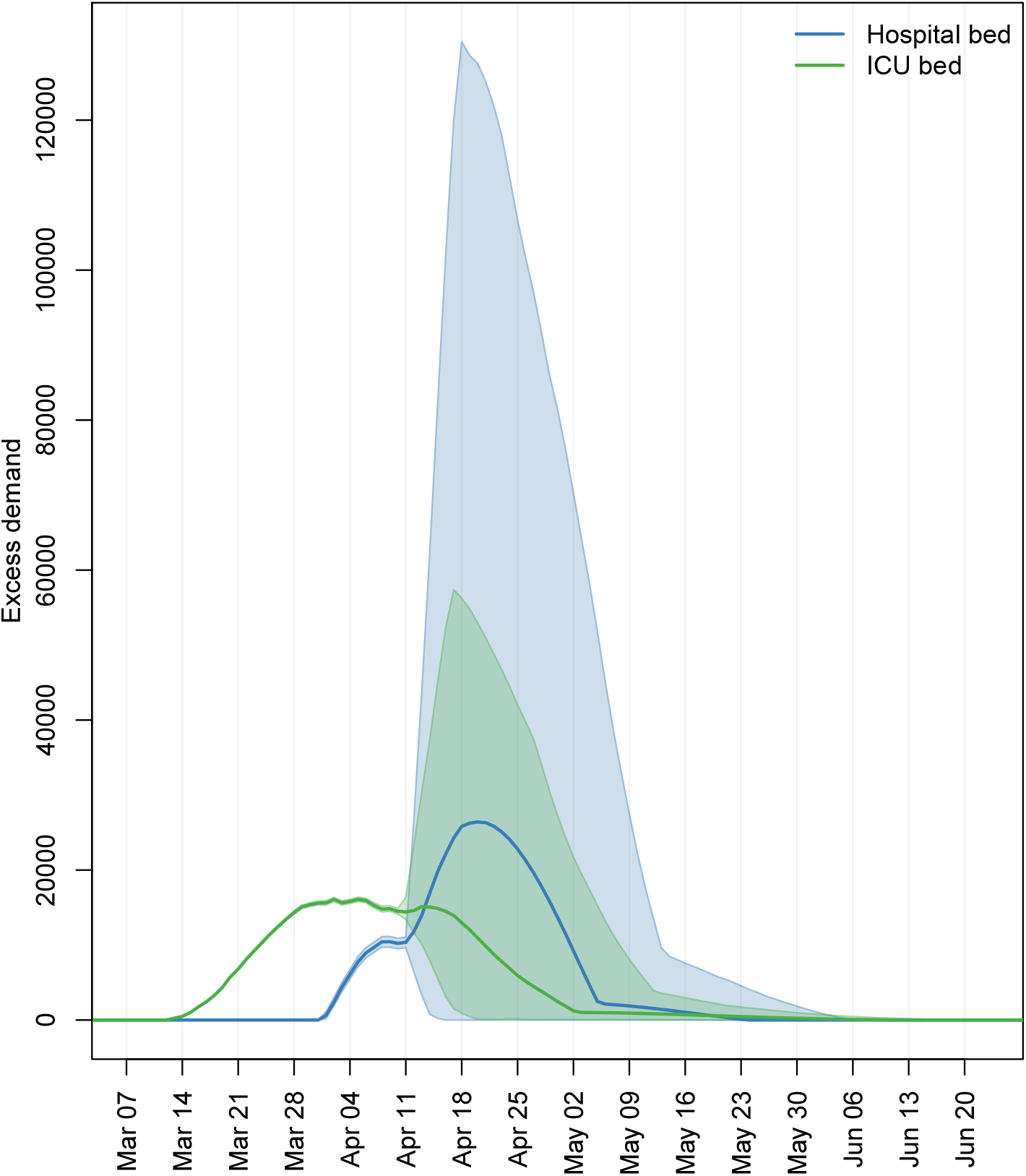
Excess demand for services above currently available capacity

**Figure 6.**
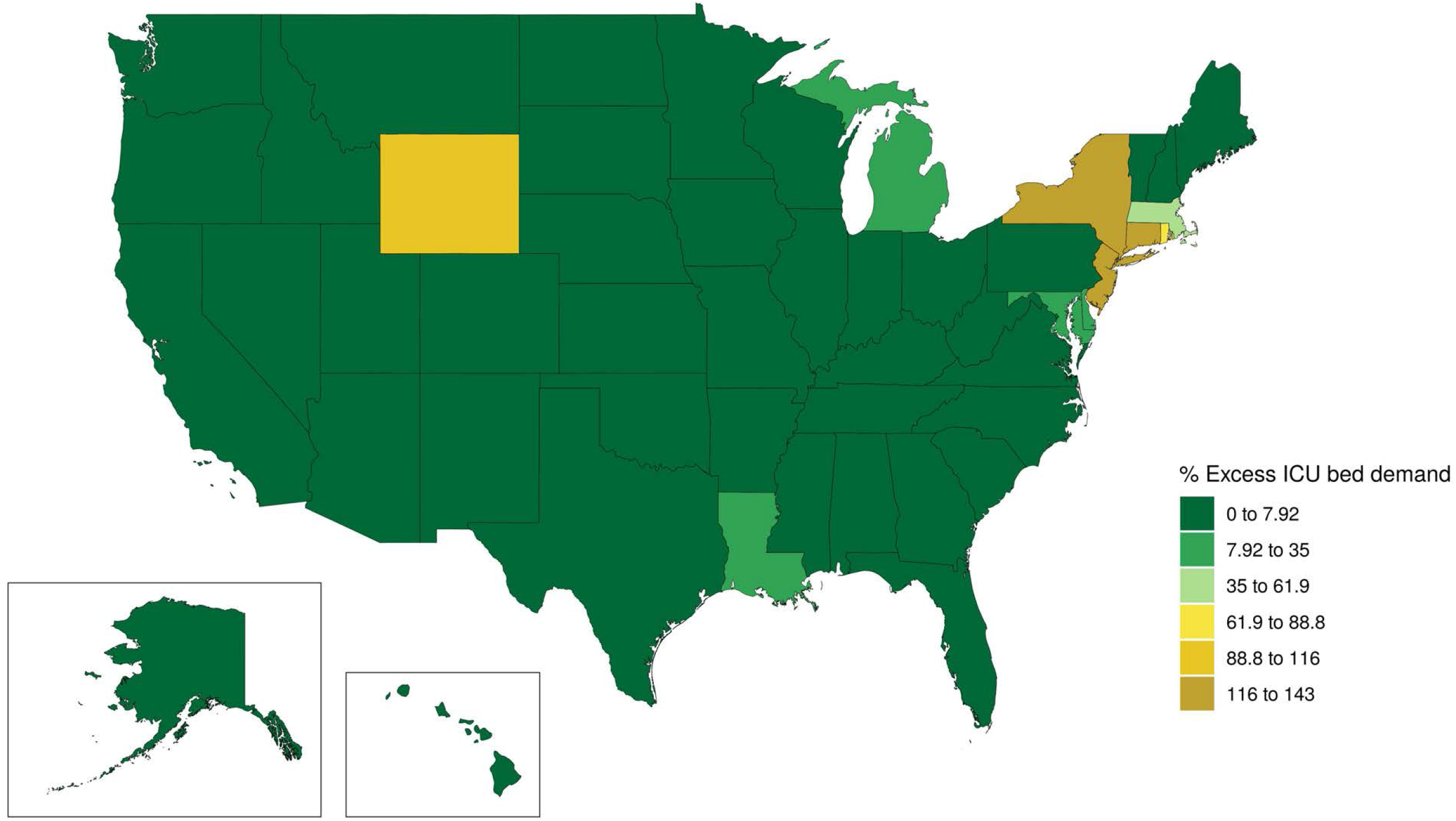
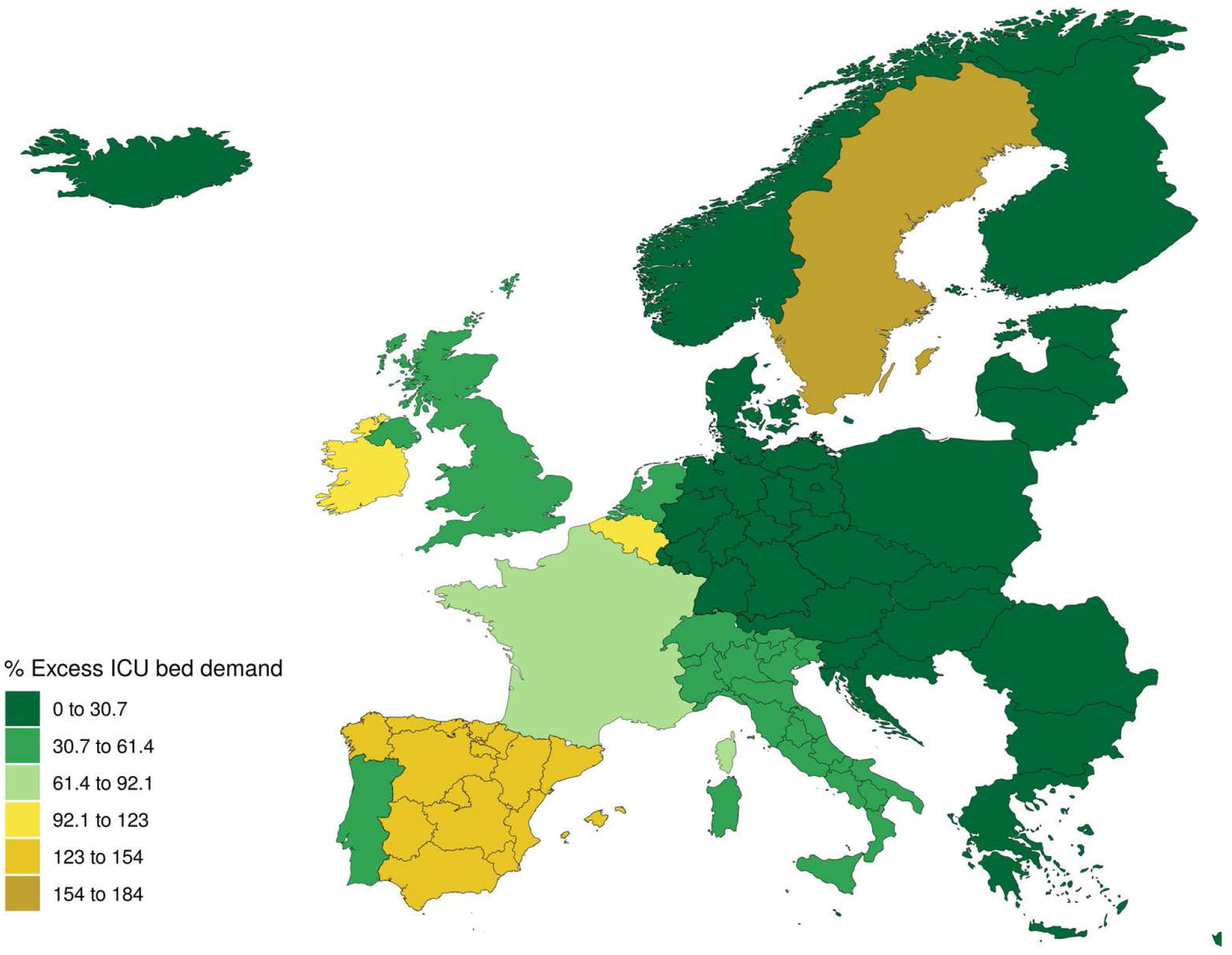
Peak percentage excess demand by location for ICU beds for the USA (Panel A) and EEA (Panel B)

Figure 7 shows the expected cumulative death numbers with 95% uncertainty intervals for the USA (Panel A) and EEA (Panel B). In the USA, the average forecast suggests 60,308 deaths, but the range is large, from 34,063 to 140,381 deaths. The figure shows that uncertainty widens markedly as the peak of the epidemic approaches, given that the exact timing of the peak is uncertain. In the EEA, 91,972 (95% UI 91,212–93,620) deaths have already been recorded so far, with the majority of these coming from Italy, Spain, and France. Our forecast suggests a cumulative total of 143,088 (101,131–253,163) deaths in the EEA. A large number of these deaths are projected to occur in the UK (13,759 observed to date; 37,521 [17,625–89,385] total), Sweden (1,333 observed to date; 5,890 [1,965–16,883] total), Germany (3,570 observed to date; 4,957 [3,697–9,379] total) and France (18,485 observed to date; 22,555 [19,455–29,314] total).

**Figure 7.**
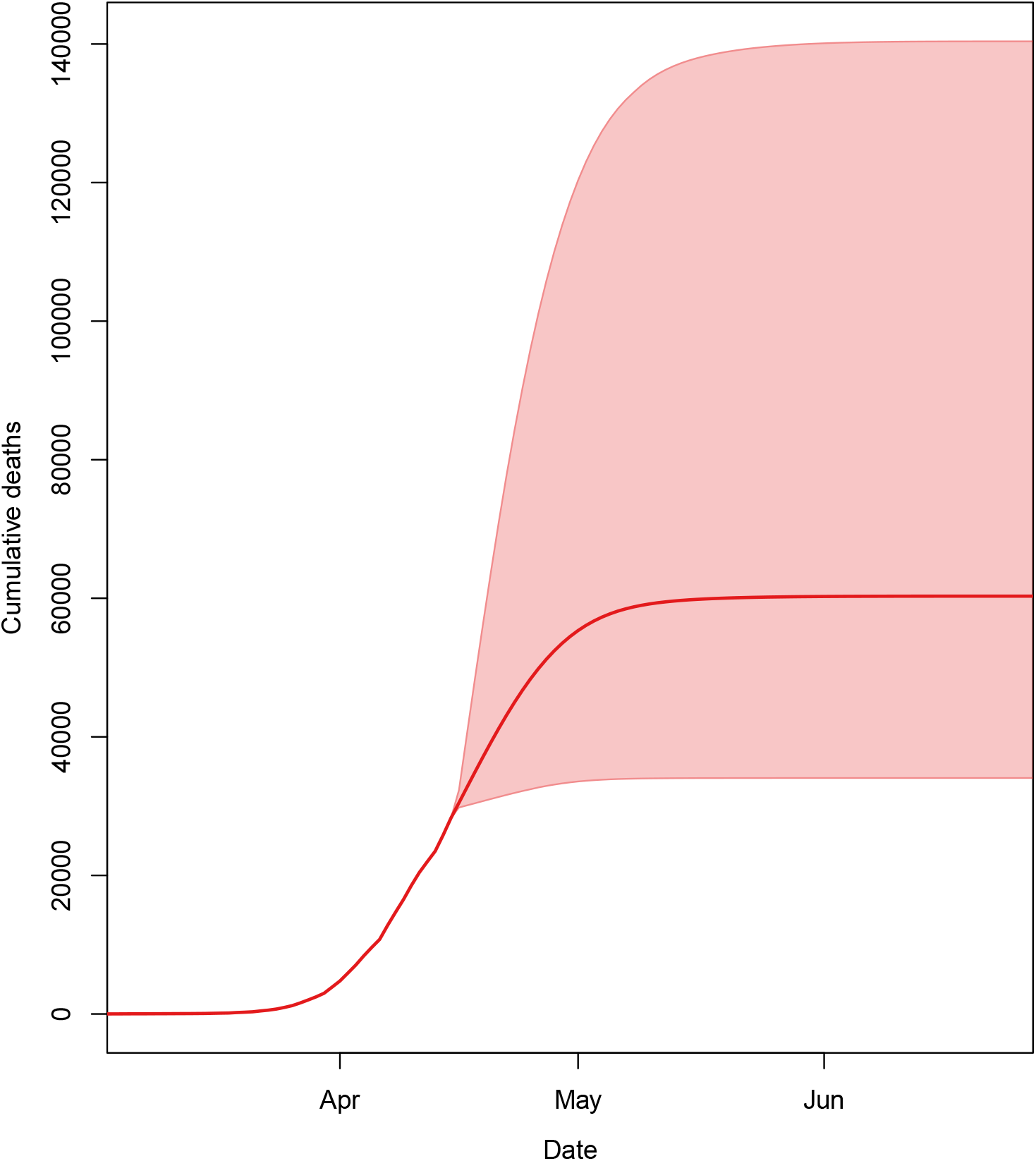
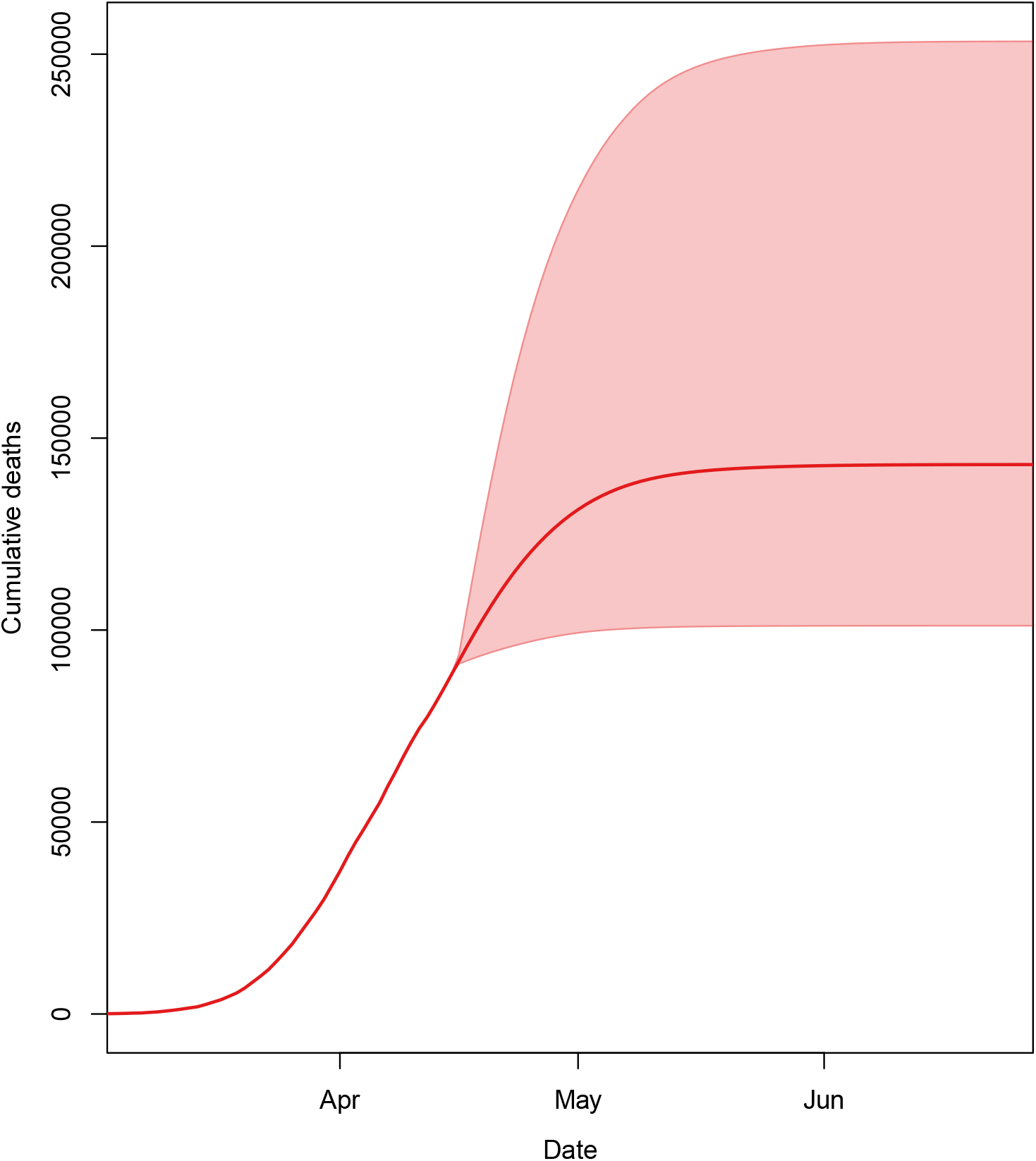
Expected cumulative death numbers with 95% uncertainty intervals for the US (Panel A) and EEA (Panel B)

Figure 8 shows a map of the cumulative number of deaths per capita by location for the USA and EEA. In Europe, the highest number of estimated cumulative deaths per capita are in Italy – particularly the northern regions – Spain, Belgium, Sweden and the UK. In the USA, states with the highest per capita deaths are New York, Rhode Island, New Jersey, Connecticut, Massachusetts, Wyoming, Louisiana, and Michigan.

**Figure 8.**
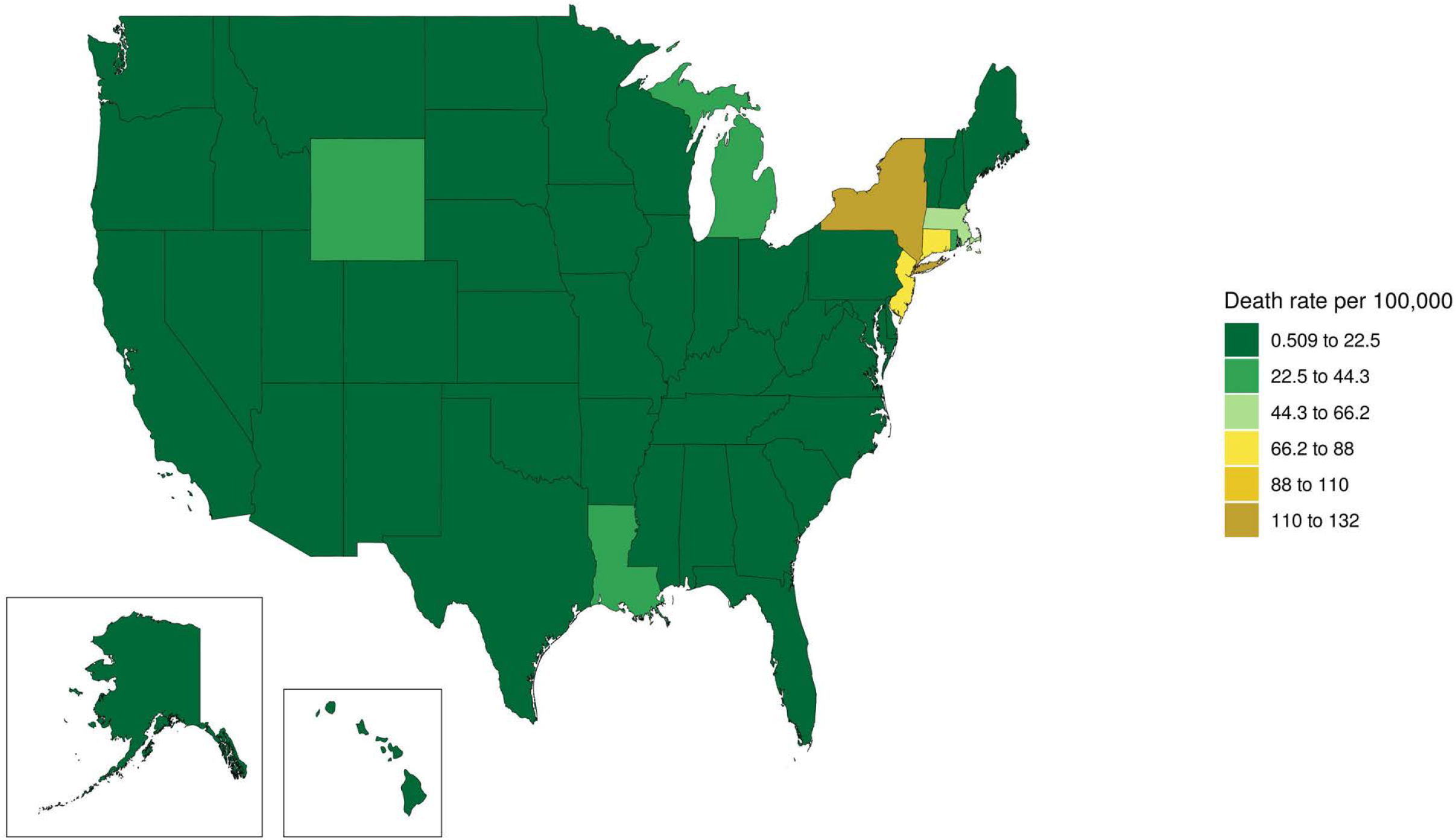
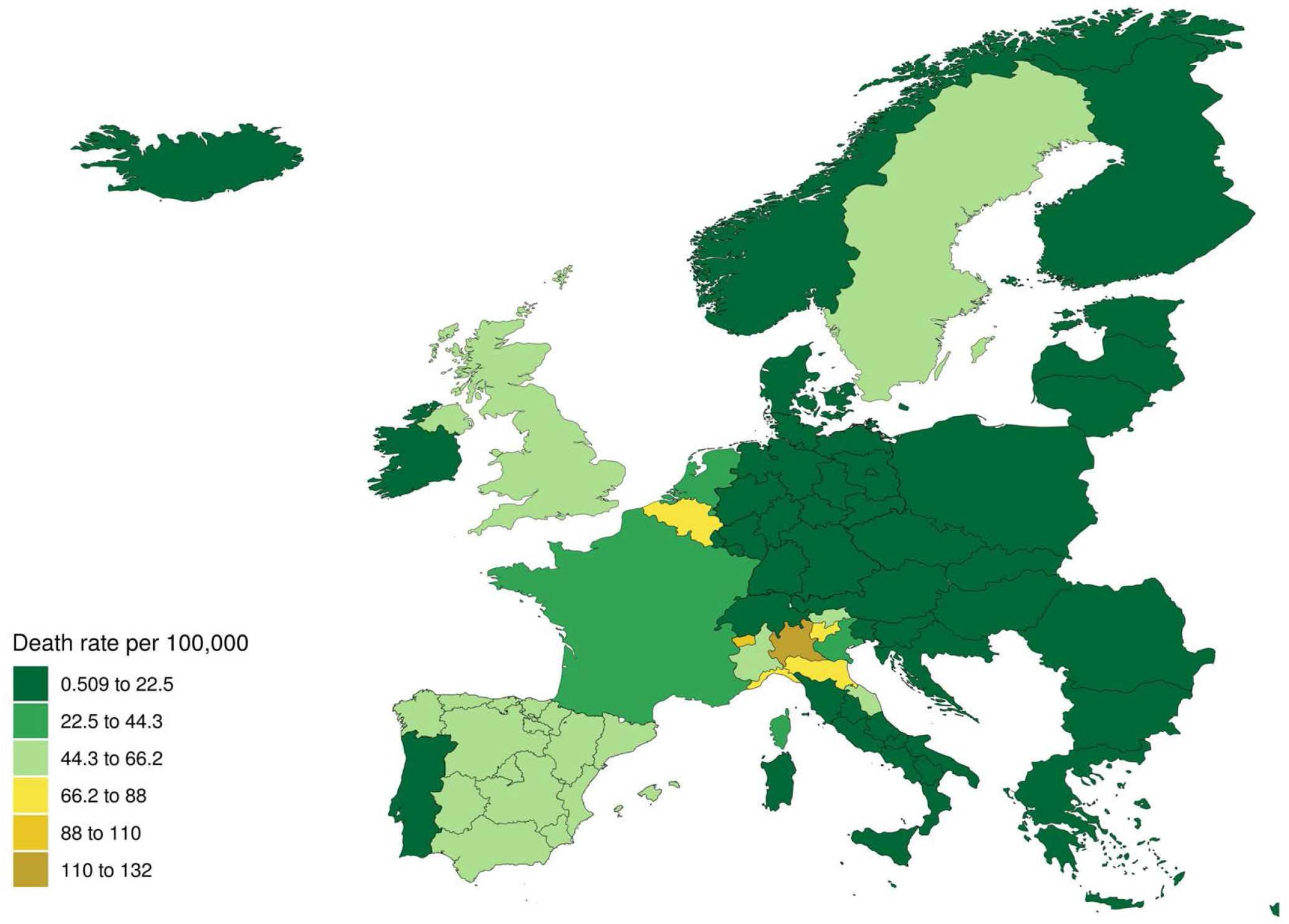
Cumulative deaths per 100,000 population for the USA (Panel A) and EEA (Panel B).

Figure 9 shows the date by location by which projected daily deaths drop below 0.3 per million. As expected, there is a strong correlation between the timing of the peak daily death and when the daily death rate will drop below this threshold. In Europe, countries where this will happen later include the UK, Norway, Denmark, Sweden and the Netherlands. In the USA, states that will not cross this threshold until the end of May include South Dakota, North Dakota, Iowa, Oklahoma, Arkansas and Utah.

**Figure 9.**
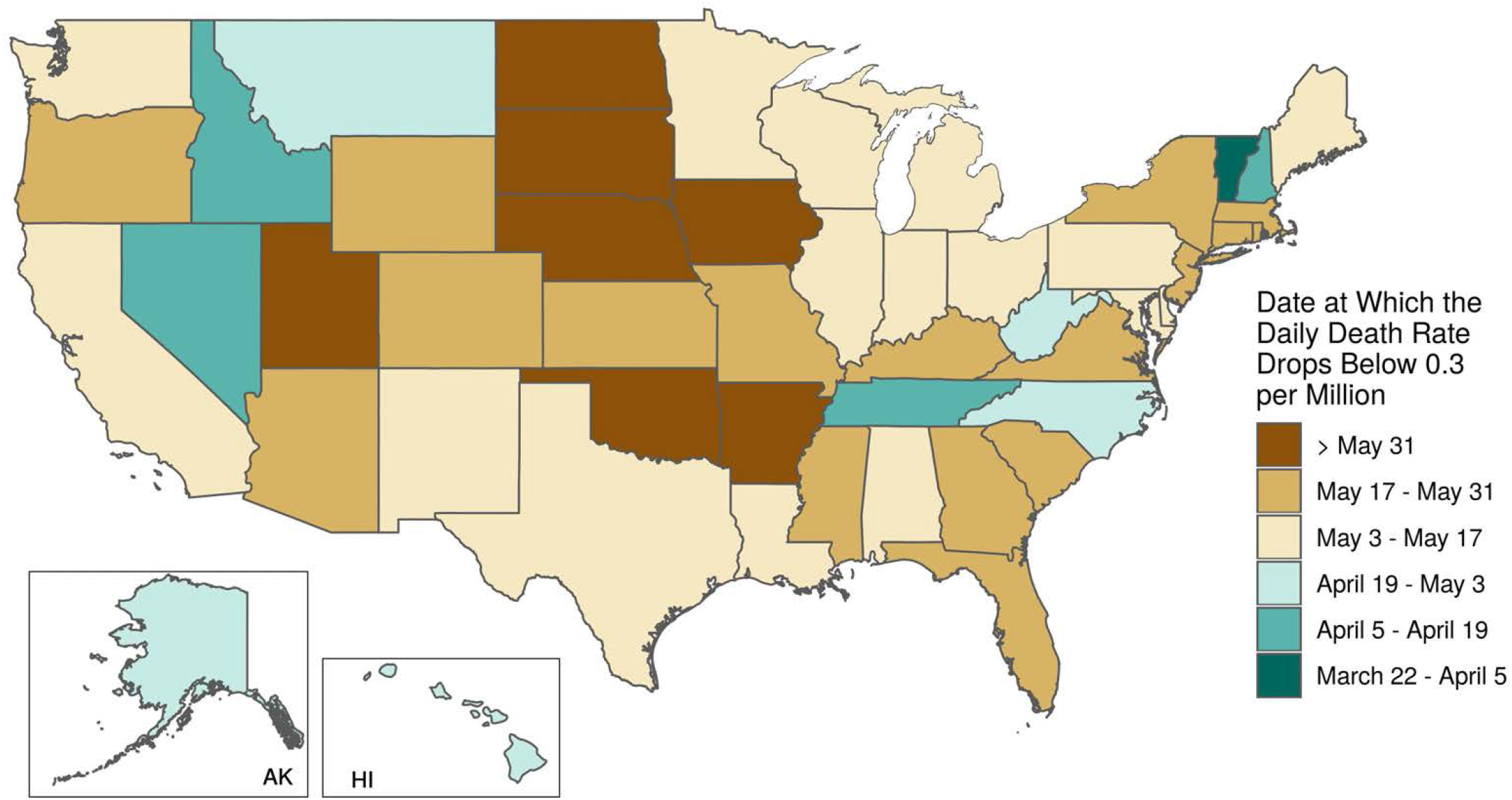
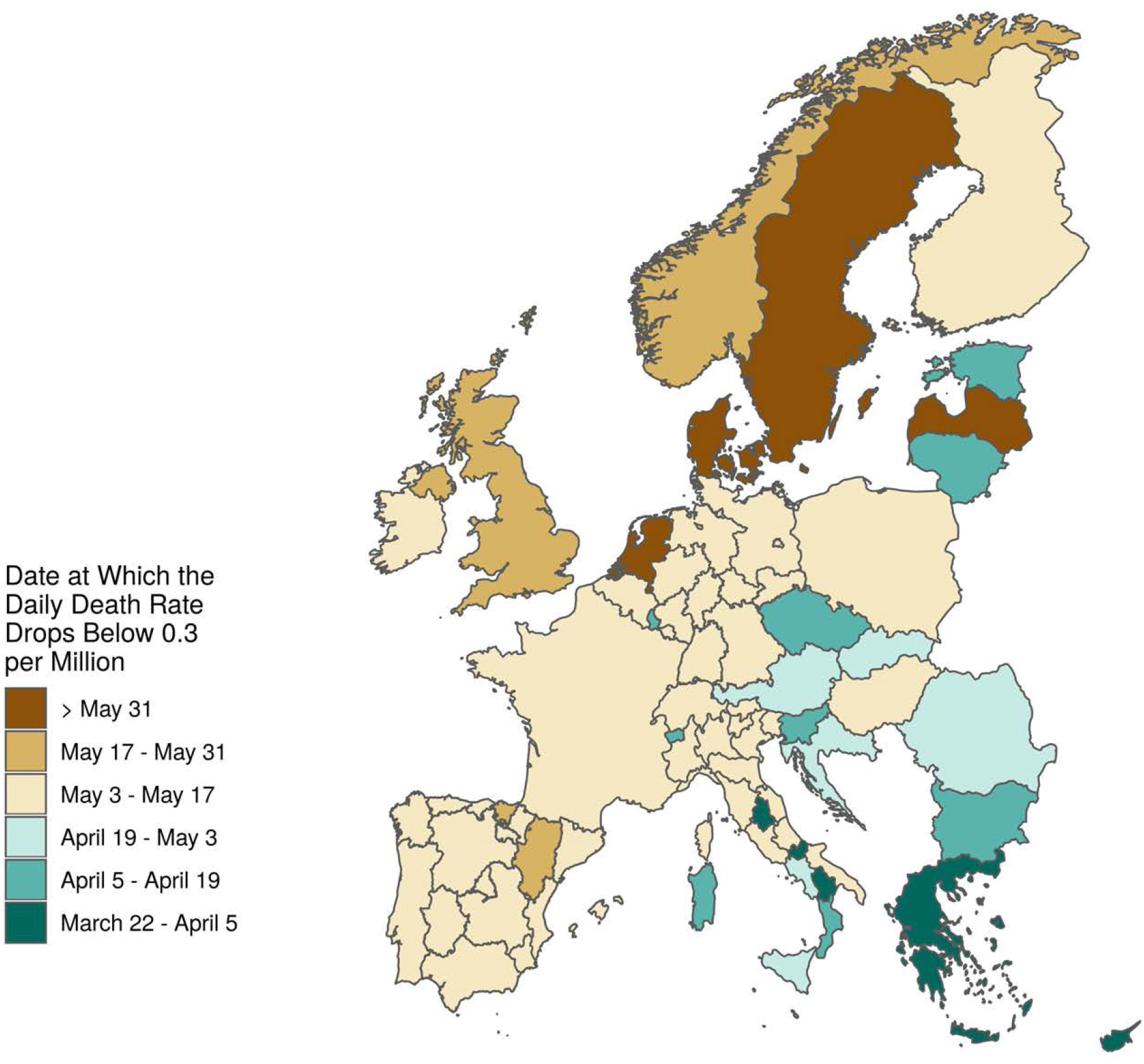
Date at which the daily death rate is projected to drop below 0.3 per million by location for the USA (Panel A) and EEA (Panel B)

Results for each location are accessible through a visualisation tool at http://covid19.healthdata.org/projections – the estimates presented in this tool will be continually updated as new data are incorporated and ultimately will supersede the results in this paper. Summary information on cumulative deaths, the date of peak demand, the peak demand, peak excess demand, and aggregate demand are provided for each location in Table 1.

**Table 1.** Summary information on deaths, peak demand, peak excess demand, and aggregate demand by location

| Location name | Cumulative Deaths | Date of Peak Hospital use | Beds Used at Peak | ICU Beds Used at Peak | Ventilators Used at Peak | Excess Bed Demand | Excess ICU Demand | Cumulative Bed Days | Cumulative ICU Days | Cumulative Ventilator Days |
| --- | --- | --- | --- | --- | --- | --- | --- | --- | --- | --- |
| Austria | 457 (414–593) | 04/06/2020 (04/06/2020–04/17/2020) | 703 (679–801) | 183 (179–220) | 165 (163–197) | 0 (0–0) | 0 (0–0) | 703 (13321–20478) | 4062 (3608–5343) | 3600 (3204–4719) |
| Belgium | 8039 (5416–15180) | 04/11/2020 (04/11/2020–04/18/2020) | 10321 (9994–27826) | 2730 (2544–7094) | 2432 (2320–6603) | 0 (0–1078) | 1931 (1746–6295) | 10321 (171751–506232) | 70826 (47069–134970) | 62930 (41880–119972) |
| Bulgaria | 47 (38–86) | 04/12/2020 (04/11/2020–04/17/2020) | 70 (61–187) | 19 (17–47) | 17 (16–44) | 0 (0–0) | 0 (0–0) | 70 (1138–3134) | 425 (315–800) | 376 (281–699) |
| Croatia | 56 (35–114) | 04/14/2020 (04/12/2020–04/18/2020) | 97 (64–280) | 25 (19–68) | 23 (17–64) | 0 (0–0) | 0 (0–0) | 97 (1044–4082) | 498 (294–1052) | 442 (262–923) |
| Cyprus | 18 (14–33) | 03/28/2020 (03/28/2020–04/18/2020) | 37 (30–83) | 8 (8–19) | 8 (7–17) | 0 (0–0) | 0 (0–0) | 37 (446–1528) | 169 (116–344) | 146 (103–298) |
| Czechia | 194 (170–274) | 04/09/2020 (04/09/2020–04/15/2020) | 358 (340–535) | 89 (86–136) | 79 (78–123) | 0 (0–0) | 0 (0–0) | 358 (5711–10170) | 1766 (1508–2541) | 1555 (1333–2228) |
| Denmark | 683 (354–1637) | 04/18/2020 (04/04/2020–04/18/2020) | 549 (520–1885) | 139 (131–438) | 124 (119–405) | 0 (0–0) | 47 (39–346) | 549 (11663–58026) | 6155 (3103–14980) | 5436 (2744–13196) |
| Estonia | 57 (36–135) | 04/03/2020 (04/03/2020–04/18/2020) | 79 (70–211) | 20 (18–50) | 18 (17–48) | 0 (0–0) | 0 (0–9) | 79 (1034–4639) | 501 (297–1214) | 445 (266–1076) |
| Finland | 118 (76–257) | 04/13/2020 (04/11/2020–04/18/2020) | 150 (137–400) | 40 (38–102) | 37 (35–94) | 0 (0–0) | 0 (0–41) | 150 (2278–8531) | 1043 (644–2279) | 926 (576–2019) |
| France | 22555 (19455–29314) | 04/02/2020 (04/02/2020–04/16/2020) | 25795 (25297–32686) | 6975 (6907–8803) | 6266 (6213–7968) | 0 (0–0) | 5214 (5146–7042) | 25795 (612478–957724) | 197059 (168591–258463) | 175462 (150150–229655) |
| Germany | 4957 (3697–9379) | 04/11/2020 (04/11/2020–04/17/2020) | 6322 (6241–16734) | 1778 (1637–4432) | 1596 (1481–4130) | 0 (0–0) | 0 (0–0) | 6322 (114421–296138) | 42929 (31823–81389) | 38317 (28433–72531) |
| Bavaria | 1431 (1086–2741) | 04/11/2020 (04/11/2020–04/17/2020) | 1809 (1760–4998) | 514 (473–1322) | 461 (428–1236) | 0 (0–0) | 0 (0–0) | 1809 (33175–87392) | 12393 (9306–23942) | 11062 (8328–21320) |
| Berlin | 159 (81–455) | 04/16/2020 (04/11/2020–04/18/2020) | 260 (147–1023) | 69 (38–259) | 64 (36–245) | 0 (0–0) | 0 (0–0) | 260 (2327–14389) | 1374 (670–3984) | 1226 (603–3521) |
| Brandenburg | 70 (54–126) | 04/15/2020 (04/05/2020–04/17/2020) | 119 (108–313) | 34 (28–80) | 31 (27–76) | 0 (0–0) | 0 (0–0) | 119 (1535–4141) | 604 (447–1104) | 539 (401–983) |
| Bremen | 26 (21–50) | 04/09/2020 (04/09/2020–04/18/2020) | 48 (42–95) | 13 (12–26) | 12 (11–24) | 0 (0–0) | 0 (0–0) | 48 (553–1654) | 230 (165–445) | 205 (149–394) |
| Hamburg | 139 (84–321) | 04/16/2020 (04/06/2020–04/18/2020) | 217 (159–606) | 60 (45–156) | 55 (42–146) | 0 (0–0) | 0 (0–0) | 217 (2433–10064) | 1205 (700–2751) | 1075 (629–2447) |
| Hesse | 290 (196–644) | 04/14/2020 (04/11/2020–04/18/2020) | 423 (360–1221) | 117 (98–315) | 106 (90–295) | 0 (0–0) | 0 (0–0) | 423 (5809–20528) | 2515 (1652–5607) | 2245 (1481–4988) |
| Lower Saxony | 326 (253–569) | 04/11/2020 (04/11/2020–04/17/2020) | 461 (441–974) | 124 (121–257) | 114 (112–235) | 0 (0–0) | 0 (0–0) | 461 (7553–18495) | 2820 (2142–5025) | 2517 (1917–4454) |
| Mecklenburg–Vorpommern | 17 (13–35) | 04/03/2020 (04/03/2020–04/17/2020) | 35 (30–79) | 9 (9–19) | 9 (8–18) | 0 (0–0) | 0 (0–0) | 35 (330–1195) | 151 (99–314) | 135 (91–279) |
| North Rhine–Westphalia | 841 (692–1239) | 04/11/2020 (04/11/2020–04/16/2020) | 1187 (1153–2098) | 321 (306–552) | 286 (281–512) | 0 (0–0) | 0 (0–0) | 1187 (21113–39503) | 7285 (5924–10804) | 6501 (5294–9643) |
| Rhineland–Palatinate | 170 (93–498) | 04/16/2020 (04/11/2020–04/18/2020) | 252 (153–1096) | 67 (44–274) | 61 (41–260) | 0 (0–0) | 0 (0–0) | 252 (2680–15767) | 1473 (774–4321) | 1315 (695–3848) |
| Saarland | 116 (71–267) | 04/11/2020 (04/11/2020–04/17/2020) | 195 (181–484) | 52 (41–122) | 48 (39–114) | 0 (0–0) | 0 (0–0) | 195 (2077–8513) | 1006 (603–2312) | 898 (540–2054) |
| Saxony–Anhalt | 36 (27–74) | 04/09/2020 (04/09/2020–04/18/2020) | 54 (47–166) | 15 (14–45) | 14 (13–42) | 0 (0–0) | 0 (0–0) | 54 (716–2464) | 315 (213–655) | 281 (193–584) |
| Saxony | 137 (94–276) | 04/16/2020 (04/11/2020–04/17/2020) | 201 (169–517) | 57 (45–131) | 52 (41–124) | 0 (0–0) | 0 (0–0) | 201 (2765–8860) | 1182 (796–2412) | 1055 (714–2152) |
| Schleswig–Holstein | 70 (55–113) | 04/10/2020 (04/10/2020–04/18/2020) | 110 (100–190) | 28 (26–53) | 25 (24–49) | 0 (0–0) | 0 (0–0) | 110 (1553–3738) | 609 (454–1002) | 543 (408–892) |
| Thuringia | 67 (45–135) | 04/14/2020 (04/11/2020–04/16/2020) | 112 (96–211) | 31 (26–54) | 29 (24–50) | 0 (0–0) | 0 (0–0) | 112 (1255–4305) | 576 (367–1161) | 514 (330–1037) |
| Greece | 119 (105–176) | 04/01/2020 (04/01/2020–04/17/2020) | 149 (137–246) | 41 (40–68) | 38 (36–63) | 0 (0–0) | 0 (0–0) | 149 (2880–5705) | 1018 (852–1566) | 912 (769–1398) |
| Hungary | 305 (153–795) | 04/18/2020 (04/10/2020–04/18/2020) | 507 (342–1907) | 129 (85–435) | 116 (79–408) | 0 (0–0) | 0 (0–0) | 507 (5076–27919) | 2757 (1357–7156) | 2433 (1199–6297) |
| Iceland | 19 (9–81) | 04/06/2020 (04/06/2020–05/04/2020) | 20 (16–72) | 5 (4–18) | 5 (4–16) | 0 (0–0) | 0 (0–5) | 20 (236–3114) | 175 (68–773) | 153 (62–676) |
| Ireland | 890 (516–2297) | 04/15/2020 (04/11/2020–04/18/2020) | 1410 (1149–5741) | 327 (271–1201) | 294 (245–1122) | 0 (0–2438) | 256 (200–1130) | 1410 (19072–90886) | 8379 (4763–22025) | 7316 (4171–19214) |
| Italy | 26007 (23589–31056) | 03/28/2020 (03/28/2020–04/17/2020) | 24029 (23651–24429) | 6681 (6630–6825) | 6089 (6046–6249) | 0 (0–0) | 4622 (4571–4766) | 24029 (702638–951530) | 222179 (199885–266815) | 199048 (179250–238960) |
| Abruzzo | 286 (255–354) | 03/29/2020 (03/29/2020–04/18/2020) | 322 (306–347) | 91 (88–102) | 84 (82–93) | 0 (0–0) | 0 (0–0) | 322 (7219–11051) | 2437 (2110–3073) | 2185 (1900–2743) |
| Basilicata | 23 (22–30) | 03/31/2020 (03/31/2020–04/15/2020) | 36 (31–43) | 10 (9–12) | 9 (9–11) | 0 (0–0) | 0 (0–0) | 36 (541–1000) | 197 (167–266) | 176 (153–237) |
| Calabria | 77 (73–94) | 03/31/2020 (03/31/2020–03/31/2020) | 130 (120–141) | 33 (32–34) | 30 (29–31) | 0 (0–0) | 0 (0–0) | 130 (2044–3196) | 665 (593–847) | 594 (535–753) |
| Campania | 316 (294–372) | 03/23/2020 (03/23/2020–04/15/2020) | 394 (374–419) | 107 (104–110) | 94 (92–97) | 0 (0–0) | 0 (0–0) | 394 (9413–13085) | 2819 (2550–3382) | 2498 (2264–2990) |
| Emilia–Romagna | 3297 (3065–3764) | 03/26/2020 (03/26/2020–03/26/2020) | 2727 (2634–2828) | 784 (769–799) | 705 (694–717) | 0 (0–0) | 0 (0–0) | 2727 (87661–114738) | 27950 (25536–32304) | 25092 (22981–28980) |
| Friuli–Venezia Giulia | 256 (230–312) | 03/29/2020 (03/29/2020–03/29/2020) | 241 (229–255) | 68 (66–70) | 62 (61–65) | 0 (0–0) | 0 (0–0) | 241 (6383–9640) | 2166 (1889–2691) | 1945 (1703–2409) |
| Lazio | 376 (335–464) | 03/30/2020 (03/30/2020–04/15/2020) | 381 (365–431) | 103 (100–120) | 93 (91–109) | 0 (0–0) | 0 (0–0) | 381 (9920–15050) | 3248 (2820–4093) | 2902 (2529–3644) |
| Liguria | 1046 (914–1290) | 03/27/2020 (03/27/2020–04/18/2020) | 799 (776–994) | 244 (240–290) | 221 (217–268) | 0 (0–0) | 0 (0–0) | 799 (24640–36922) | 8677 (7451–10830) | 7838 (6743–9778) |
| Lombardia | 13162 (12220–15146) | 03/27/2020 (03/27/2020–03/27/2020) | 13311 (12956–13654) | 3715 (3666–3767) | 3371 (3332–3412) | 0 (0–0) | 0 (0–0) | 13311 (363813–476190) | 112990 (103300–131753) | 101090 (92642–117739) |
| Marche | 879 (804–1026) | 03/29/2020 (03/29/2020–03/29/2020) | 904 (876–935) | 260 (256–265) | 237 (233–240) | 0 (0–0) | 0 (0–0) | 904 (22598–30732) | 7407 (6652–8749) | 6660 (6001–7853) |
| Molise | 16 (16–16) | 03/19/2020 (03/19/2020–03/19/2020) | 28 (24–33) | 8 (8–9) | 8 (7–8) | 0 (0–0) | 0 (0–0) | 28 (367–605) | 135 (117–158) | 121 (108–139) |
| Piemonte | 2882 (2365–3876) | 04/12/2020 (04/11/2020–04/18/2020) | 2665 (2570–4430) | 746 (734–1242) | 681 (673–1142) | 0 (0–0) | 0 (0–0) | 2665 (67471–116914) | 24351 (19743–33335) | 21881 (17756–29822) |
| Provincia autonoma di Bolzano | 273 (237–337) | 03/31/2020 (03/31/2020–03/31/2020) | 409 (390–428) | 113 (111–116) | 104 (102–106) | 0 (0–0) | 0 (0–0) | 409 (7129–11229) | 2382 (2005–2996) | 2121 (1792–2663) |
| Provincia autonoma di Trento | 402 (341–556) | 03/26/2020 (03/26/2020–04/18/2020) | 448 (429–627) | 116 (113–172) | 105 (103–160) | 0 (0–0) | 0 (0–0) | 448 (10092–17737) | 3471 (2873–4860) | 3101 (2575–4336) |
| Puglia | 383 (330–480) | 04/01/2020 (04/01/2020–04/17/2020) | 440 (421–467) | 115 (113–126) | 107 (105–114) | 0 (0–0) | 0 (0–0) | 440 (9882–15658) | 3320 (2787–4238) | 2962 (2500–3766) |
| Sardegna | 92 (85–120) | 04/06/2020 (04/06/2020–04/15/2020) | 140 (130–165) | 38 (37–46) | 35 (34–42) | 0 (0–0) | 0 (0–0) | 140 (2408–4014) | 799 (696–1073) | 714 (626–953) |
| Sicilia | 208 (192–252) | 03/27/2020 (03/27/2020–03/27/2020) | 269 (252–285) | 69 (67–78) | 64 (62–70) | 0 (0–0) | 0 (0–0) | 269 (5727–8398) | 1817 (1615–2242) | 1619 (1447–1993) |
| Toscana | 724 (633–913) | 04/11/2020 (04/11/2020–04/17/2020) | 701 (677–994) | 198 (194–281) | 180 (178–258) | 0 (0–0) | 0 (0–0) | 701 (17870–27400) | 6114 (5251–7804) | 5496 (4735–7006) |
| Umbria | 55 (55–60) | 03/26/2020 (03/26/2020–03/26/2020) | 75 (67–83) | 23 (22–24) | 21 (20–22) | 0 (0–0) | 0 (0–0) | 75 (1398–1951) | 466 (429–532) | 420 (390–475) |
| Valle d'Aosta | 135 (122–178) | 03/31/2020 (03/31/2020–03/31/2020) | 198 (185–211) | 55 (53–57) | 50 (49–52) | 0 (0–0) | 0 (0–0) | 198 (3392–5682) | 1156 (999–1557) | 1035 (900–1389) |
| Veneto | 1118 (1001–1415) | 03/26/2020 (03/26/2020–04/16/2020) | 1084 (1051–1465) | 298 (293–403) | 270 (266–369) | 0 (0–0) | 0 (0–0) | 1084 (29857–44885) | 9613 (8477–12327) | 8596 (7601–10997) |
| Latvia | 80 (11–285) | 04/26/2020 (04/07/2020–04/25/2020) | 91 (14–330) | 24 (4–87) | 22 (4–79) | 0 (0–0) | 0 (0–23) | 91 (162–10786) | 705 (53–2882) | 626 (45–2559) |
| Lithuania | 39 (32–69) | 04/10/2020 (04/10/2020–04/17/2020) | 68 (60–140) | 17 (17–37) | 16 (16–34) | 0 (0–0) | 0 (0–0) | 68 (900–2367) | 345 (260–617) | 307 (234–544) |
| Luxembourg | 116 (71–265) | 04/06/2020 (04/06/2020–04/18/2020) | 153 (140–487) | 35 (34–106) | 32 (31–99) | 0 (0–0) | 0 (0–70) | 153 (2322–9997) | 1073 (624–2469) | 941 (550–2173) |
| Netherlands | 6814 (4035–14051) | 04/16/2020 (04/03/2020–04/18/2020) | 5761 (4908–15311) | 1452 (1269–3569) | 1303 (1133–3339) | 0 (0–0) | 533 (350–2650) | 5761 (133187–501154) | 61396 (35169–129013) | 54221 (31230–114002) |
| Norway | 280 (167–624) | 04/07/2020 (04/07/2020–04/18/2020) | 282 (265–828) | 73 (71–194) | 66 (64–180) | 0 (0–0) | 0 (0–93) | 282 (5510–23323) | 2547 (1462–5904) | 2243 (1294–5206) |
| Portugal | 980 (661–2003) | 04/16/2020 (04/11/2020–04/18/2020) | 1096 (952–3928) | 302 (266–996) | 271 (240–940) | 0 (0–0) | 190 (154–884) | 1096 (20181–63950) | 8507 (5650–17513) | 7588 (5050–15593) |
| Poland | 646 (337–2020) | 04/16/2020 (04/11/2020–04/18/2020) | 771 (678–3389) | 196 (180–753) | 174 (163–702) | 0 (0–0) | 0 (0–0) | 771 (11554–72641) | 5904 (3011–18409) | 5195 (2656–16211) |
| Romania | 618 (413–1489) | 04/16/2020 (04/07/2020–04/18/2020) | 923 (749–4173) | 241 (188–1034) | 215 (170–973) | 0 (0–0) | 0 (0–0) | 923 (13782–53650) | 5561 (3656–13879) | 4913 (3233–12219) |
| Slovakia | 252 (41–955) | 04/18/2020 (04/19/2020–04/18/2020) | 1123 (149–4409) | 237 (33–935) | 226 (30–886) | 0 (0–0) | 49 (0–747) | 1123 (1506–37134) | 2374 (378–9002) | 2072 (331–7865) |
| Slovenia | 70 (61–105) | 04/06/2020 (04/06/2020–04/18/2020) | 124 (113–177) | 30 (29–48) | 28 (27–44) | 0 (0–0) | 0 (0–8) | 124 (1803–3718) | 622 (507–951) | 552 (454–840) |
| Spain | 23680 (20269–31608) | 03/29/2020 (03/29/2020–04/17/2020) | 26474 (26066–35145) | 6961 (6905–9398) | 6343 (6302–8652) | 0 (0–3355) | 5597 (5541–8034) | 26474 (637743–1018635) | 206496 (175544–276670) | 183962 (156513–246374) |
| Andalucia | 1099 (950–1492) | 04/06/2020 (04/06/2020–04/16/2020) | 1486 (1445–1900) | 414 (408–515) | 377 (372–470) | 0 (0–0) | 0 (0–0) | 1486 (29342–48738) | 9583 (8150–13152) | 8539 (7278–11707) |
| Aragon | 759 (605–1056) | 04/04/2020 (04/04/2020–04/16/2020) | 815 (786–1182) | 234 (230–318) | 210 (207–289) | 0 (0–0) | 0 (0–0) | 815 (18687–34310) | 6620 (5204–9286) | 5897 (4642–8263) |
| Asturias | 225 (172–410) | 04/09/2020 (04/09/2020–04/17/2020) | 284 (269–768) | 75 (73–201) | 68 (67–189) | 0 (0–0) | 0 (0–0) | 284 (5126–13353) | 1966 (1455–3556) | 1752 (1301–3175) |
| Balearic Islands | 154 (131–228) | 03/28/2020 (03/28/2020–04/16/2020) | 203 (190–333) | 53 (51–88) | 49 (47–81) | 0 (0–0) | 0 (0–0) | 203 (3868–7619) | 1340 (1099–2016) | 1194 (986–1796) |
| Canary Islands | 115 (107–134) | 03/28/2020 (03/28/2020–03/28/2020) | 195 (181–211) | 53 (52–55) | 49 (47–50) | 0 (0–0) | 0 (0–0) | 195 (3115–4594) | 999 (888–1209) | 890 (797–1071) |
| Cantabria | 173 (137–295) | 03/31/2020 (03/31/2020–04/17/2020) | 212 (198–475) | 55 (53–124) | 49 (48–115) | 0 (0–0) | 0 (0–0) | 212 (4080–9799) | 1507 (1158–2616) | 1342 (1035–2324) |
| Castilla–La Mancha | 2475 (1959–3630) | 04/08/2020 (04/08/2020–04/17/2020) | 2432 (2362–4882) | 656 (646–1251) | 587 (579–1161) | 0 (0–0) | 0 (0–0) | 2432 (60936–118300) | 21584 (16887–31923) | 19230 (15077–28405) |
| Catalonia | 4743 (4100–6320) | 03/29/2020 (03/28/2020–04/17/2020) | 6281 (6028–6928) | 1657 (1626–1826) | 1526 (1502–1687) | 0 (0–0) | 0 (0–0) | 6281 (125495–206956) | 41367 (35004–55711) | 36852 (31290–49585) |
| Community of Madrid | 7867 (7181–9195) | 03/24/2020 (03/24/2020–03/24/2020) | 9182 (8895–9492) | 2398 (2358–2440) | 2176 (2147–2210) | 0 (0–0) | 0 (0–0) | 9182 (221673–303231) | 68614 (61563–81483) | 61125 (54960–72519) |
| Extremadura | 468 (378–700) | 03/28/2020 (03/28/2020–04/17/2020) | 561 (539–931) | 148 (145–244) | 135 (133–226) | 0 (0–0) | 0 (0–0) | 561 (11587–22901) | 4083 (3236–6173) | 3637 (2889–5493) |
| Galicia | 378 (324–518) | 03/31/2020 (03/31/2020–04/16/2020) | 530 (507–615) | 133 (130–163) | 124 (122–149) | 0 (0–0) | 0 (0–0) | 530 (9859–17020) | 3296 (2767–4562) | 2936 (2474–4052) |
| La Rioja | 345 (279–515) | 04/06/2020 (04/06/2020–04/17/2020) | 384 (365–640) | 105 (102–171) | 95 (93–156) | 0 (0–0) | 0 (0–0) | 384 (8494–17097) | 3013 (2383–4593) | 2684 (2130–4073) |
| Murcia | 129 (111–196) | 04/02/2020 (04/02/2020–04/17/2020) | 210 (197–308) | 56 (54–82) | 51 (49–77) | 0 (0–0) | 0 (0–0) | 210 (3270–6519) | 1123 (929–1727) | 1000 (832–1539) |
| Navarre | 319 (270–469) | 03/29/2020 (03/29/2020–04/16/2020) | 436 (416–647) | 113 (111–167) | 104 (102–156) | 0 (0–0) | 0 (0–0) | 436 (8166–15535) | 2778 (2290–4171) | 2475 (2049–3705) |
| Valencian Community | 1220 (1016–1762) | 03/30/2020 (03/30/2020–04/17/2020) | 1505 (1461–2394) | 395 (389–630) | 361 (356–585) | 0 (0–0) | 0 (0–0) | 1505 (31510–57416) | 10640 (8738–15508) | 9480 (7803–13803) |
| Sweden | 5890 (1965–16883) | 04/29/2020 (04/11/2020–04/21/2020) | 4173 (2350–13825) | 1099 (652–3672) | 979 (577–3235) | 2365 (542–12017) | 1020 (573–3593) | 4173 (62421–568761) | 52143 (17091–151841) | 46270 (15174–134608) |
| United Kingdom | 37521 (17625–89385) | 04/20/2020 (04/08/2020–04/18/2020) | 42407 (27463–132942) | 10646 (6947–31240) | 9577 (6312–28710) | 24642 (9698–115177) | 3865 (166–24459) | 42407 (600161–3074135) | 337062 (157746–800822) | 297904 (139414–707443) |
| United States of America | 60308 (34063–140381) | 04/15/2020 (04/11/2020–04/18/2020) | 68884 (57922–226051) | 18286 (15494–54755) | 16631 (14188–51508) | 9079 (3857–88921) | 9356 (7718–38347) | 68884 (1049489–4558549) | 523419 (292547–1228976) | 466888 (261704–1093763) |
| Alabama | 295 (145–802) | 04/17/2020 (04/11/2020–04/18/2020) | 329 (219–1245) | 96 (64–350) | 89 (60–331) | 0 (0–0) | 0 (0–0) | 329 (3706–21985) | 2421 (1167–6624) | 2193 (1061–6010) |
| Arizona | 267 (158–682) | 04/10/2020 (04/10/2020–04/18/2020) | 312 (294–1061) | 77 (74–237) | 69 (67–220) | 0 (0–0) | 0 (0–0) | 312 (5395–26079) | 2454 (1403–6486) | 2155 (1239–5699) |
| Arkansas | 158 (37–527) | 04/30/2020 (04/12/2020–05/02/2020) | 129 (74–519) | 32 (20–128) | 28 (19–112) | 0 (0–0) | 0 (0–0) | 129 (1188–19809) | 1453 (320–4942) | 1276 (283–4332) |
| California | 1658 (1068–3548) | 04/14/2020 (04/11/2020–04/18/2020) | 2753 (2240–8233) | 633 (516–1741) | 563 (465–1608) | 0 (0–0) | 0 (0–0) | 2753 (41690–144832) | 15911 (10103–34295) | 13827 (8795–29771) |
| Colorado | 715 (389–1944) | 04/17/2020 (04/11/2020–04/18/2020) | 842 (799–3677) | 205 (193–786) | 181 (172–739) | 0 (0–0) | 0 (0–232) | 842 (14205–73527) | 6656 (3586–18172) | 5828 (3149–15942) |
| Connecticut | 2732 (1163–8601) | 04/16/2020 (04/11/2020–04/18/2020) | 3886 (2737–16784) | 935 (639–3798) | 860 (596–3504) | 2148 (999–15046) | 836 (540–3699) | 3886 (39775–312943) | 24819 (10391–79660) | 21865 (9160–70188) |
| Delaware | 143 (64–404) | 04/17/2020 (04/11/2020–04/19/2020) | 260 (162–954) | 57 (31–196) | 50 (29–178) | 0 (0–258) | 16 (0–155) | 260 (2415–17243) | 1392 (589–3993) | 1205 (514–3451) |
| District of Columbia | 170 (87–424) | 04/15/2020 (04/11/2020–04/18/2020) | 290 (238–918) | 68 (53–197) | 61 (49–180) | 0 (0–0) | 2 (0–131) | 290 (3093–17063) | 1607 (795–4112) | 1402 (698–3572) |
| Florida | 1363 (775–3430) | 04/14/2020 (04/11/2020–04/18/2020) | 1535 (1386–5229) | 405 (362–1251) | 367 (332–1180) | 0 (0–0) | 0 (0–0) | 1535 (24580–113743) | 12021 (6705–30351) | 10675 (5966–26980) |
| Georgia | 1369 (670–3828) | 04/15/2020 (04/05/2020–04/18/2020) | 1358 (1079–5783) | 350 (287–1291) | 311 (262–1209) | 0 (0–0) | 0 (0–702) | 1358 (22705–140596) | 12398 (5967–36009) | 10936 (5278–31752) |
| Hawaii | 38 (10–121) | 04/18/2020 (04/01/2020–04/18/2020) | 116 (20–434) | 26 (6–100) | 25 (5–95) | 0 (0–0) | 0 (0–55) | 116 (294–4448) | 353 (84–1122) | 310 (75–987) |
| Idaho | 63 (41–145) | 04/10/2020 (04/10/2020–04/18/2020) | 119 (109–309) | 29 (25–73) | 26 (24–67) | 0 (0–0) | 0 (0–0) | 119 (1354–5481) | 583 (358–1357) | 511 (316–1184) |
| Illinois | 2259 (1212–5054) | 04/17/2020 (04/11/2020–04/18/2020) | 3459 (2672–11295) | 837 (638–2464) | 752 (581–2310) | 0 (0–0) | 0 (0–1333) | 3459 (43782–190312) | 20895 (11093–47173) | 18327 (9744–41412) |
| Indiana | 903 (519–2529) | 04/15/2020 (04/10/2020–04/18/2020) | 1374 (1134–5590) | 329 (264–1212) | 294 (237–1130) | 0 (0–0) | 0 (0–506) | 1374 (18792–94944) | 8390 (4762–23466) | 7349 (4176–20542) |
| Iowa | 624 (106–2603) | 05/07/2020 (04/11/2020–05/08/2020) | 580 (137–2723) | 148 (35–699) | 132 (33–621) | 0 (0–0) | 0 (0–453) | 580 (3537–93259) | 5634 (942–24278) | 4973 (830–21439) |
| Kansas | 187 (88–500) | 04/17/2020 (04/09/2020–04/18/2020) | 250 (207–843) | 58 (51–180) | 51 (46–160) | 0 (0–0) | 0 (0–0) | 250 (3307–20908) | 1824 (810–4860) | 1579 (709–4199) |
| Kentucky | 407 (160–1213) | 04/21/2020 (04/08/2020–04/21/2020) | 455 (313–1806) | 110 (70–421) | 97 (62–378) | 0 (0–0) | 0 (0–0) | 455 (5660–46733) | 3789 (1455–11540) | 3318 (1275–10092) |
| Louisiana | 1685 (1269–2767) | 04/14/2020 (04/11/2020–04/18/2020) | 2619 (2355–5469) | 648 (563–1265) | 576 (505–1154) | 0 (0–0) | 171 (86–788) | 2619 (47125–107918) | 15783 (11750–26198) | 13798 (10287–22824) |
| Maine | 51 (27–134) | 04/16/2020 (04/11/2020–04/19/2020) | 83 (53–274) | 19 (13–58) | 17 (12–53) | 0 (0–0) | 0 (0–0) | 83 (916–5449) | 486 (235–1300) | 424 (207–1131) |
| Maryland | 914 (373–3160) | 04/18/2020 (04/09/2020–04/18/2020) | 2405 (1455–11111) | 441 (258–1857) | 376 (228–1629) | 0 (0–7150) | 175 (0–1591) | 2405 (20439–178003) | 10363 (4145–35424) | 8658 (3467–29604) |
| Massachusetts | 3236 (1289–9426) | 04/18/2020 (04/10/2020–04/18/2020) | 2830 (1967–10962) | 964 (685–3581) | 906 (656–3459) | 0 (0–6114) | 687 (408–3304) | 2830 (27062–211980) | 24872 (9684–73469) | 22963 (8946–67939) |
| Michigan | 3304 (2131–6780) | 04/10/2020 (04/10/2020–04/18/2020) | 4748 (4615–14655) | 1158 (1139–3347) | 1048 (1033–3110) | 0 (0–4501) | 416 (397–2605) | 4748 (74403–250353) | 30192 (19234–63001) | 26566 (16956–55325) |
| Minnesota | 195 (95–605) | 04/18/2020 (04/10/2020–04/18/2020) | 293 (230–1350) | 69 (53–278) | 62 (48–259) | 0 (0–0) | 0 (0–0) | 293 (3379–23752) | 1838 (855–5748) | 1604 (751–5002) |
| Mississippi | 369 (150–1298) | 04/23/2020 (04/06/2020–04/21/2020) | 410 (271–1985) | 102 (64–482) | 90 (58–428) | 0 (0–0) | 0 (0–142) | 410 (5225–49082) | 3421 (1349–12188) | 2999 (1186–10683) |
| Missouri | 362 (188–1027) | 04/15/2020 (04/09/2020–04/18/2020) | 474 (382–1913) | 120 (94–424) | 107 (85–397) | 0 (0–0) | 0 (0–0) | 474 (6508–37351) | 3322 (1694–9446) | 2919 (1493–8278) |
| Montana | 17 (8–43) | 04/17/2020 (03/30/2020–04/17/2020) | 40 (24–163) | 10 (7–39) | 9 (6–37) | 0 (0–0) | 0 (0–0) | 40 (224–1671) | 152 (63–414) | 134 (56–367) |
| Nebraska | 127 (21–479) | 05/01/2020 (04/06/2020–04/29/2020) | 101 (46–438) | 25 (11–110) | 22 (10–98) | 0 (0–0) | 0 (0–0) | 101 (644–17975) | 1169 (178–4524) | 1027 (158–3971) |
| Nevada | 257 (149–562) | 04/07/2020 (04/07/2020–04/18/2020) | 364 (344–1219) | 87 (83–256) | 79 (77–237) | 0 (0–0) | 0 (0–73) | 364 (5554–23196) | 2455 (1372–5511) | 2135 (1198–4795) |
| New Hampshire | 55 (32–127) | 04/15/2020 (04/10/2020–04/18/2020) | 95 (79–296) | 24 (18–68) | 21 (17–63) | 0 (0–0) | 0 (0–0) | 95 (1033–4793) | 503 (276–1186) | 442 (245–1035) |
| New Jersey | 6952 (4160–14367) | 04/15/2020 (04/11/2020–04/18/2020) | 10480 (8644–27673) | 2568 (2078–6284) | 2324 (1902–5855) | 2665 (829–19858) | 2103 (1614–5819) | 10480 (146278–527991) | 63655 (37795–133132) | 55973 (33286–117091) |
| New Mexico | 80 (36–229) | 04/18/2020 (04/10/2020–04/18/2020) | 118 (102–458) | 29 (25–104) | 26 (23–96) | 0 (0–0) | 0 (0–0) | 118 (1169–8794) | 739 (314–2196) | 649 (278–1926) |
| New York | 21812 (13623–42798) | 04/15/2020 (04/10/2020–04/17/2020) | 17346 (16061–49532) | 6039 (5567–16551) | 5603 (5208–15854) | 4336 (3051–36522) | 5321 (4849–15833) | 17346 (295917–954519) | 167923 (104084–329040) | 154958 (96145–303878) |
| North Carolina | 251 (156–529) | 04/15/2020 (04/11/2020–04/18/2020) | 523 (368–1366) | 123 (90–304) | 112 (81–281) | 0 (0–0) | 0 (0–0) | 523 (5585–20350) | 2342 (1420–4965) | 2050 (1247–4337) |
| North Dakota | 149 (9–652) | 05/04/2020 (04/10/2020–05/06/2020) | 137 (20–630) | 34 (5–157) | 30 (4–138) | 0 (0–0) | 0 (0–71) | 137 (255–24256) | 1375 (73–6065) | 1207 (65–5327) |
| Ohio | 716 (429–1645) | 04/15/2020 (04/11/2020–04/18/2020) | 1230 (934–3759) | 300 (226–847) | 274 (207–794) | 0 (0–0) | 0 (0–0) | 1230 (14924–60064) | 6547 (3863–15215) | 5760 (3401–13374) |
| Oklahoma | 359 (149–1166) | 04/15/2020 (04/06/2020–04/20/2020) | 359 (250–1396) | 86 (64–335) | 77 (58–299) | 0 (0–0) | 0 (0–0) | 359 (5161–45818) | 3331 (1333–11331) | 2919 (1174–9939) |
| Oregon | 131 (69–318) | 04/17/2020 (04/08/2020–04/18/2020) | 173 (145–603) | 38 (33–116) | 33 (30–105) | 0 (0–0) | 0 (0–0) | 173 (2815–14748) | 1326 (662–3262) | 1138 (571–2797) |
| Pennsylvania | 1707 (914–4555) | 04/15/2020 (04/11/2020–04/18/2020) | 3926 (3359–14724) | 773 (612–2512) | 663 (549–2291) | 0 (0–329) | 0 (0–1469) | 3926 (45991–236824) | 18443 (9717–49122) | 15578 (8226–41512) |
| Rhode Island | 438 (121–1656) | 04/23/2020 (04/09/2020–04/24/2020) | 610 (224–2628) | 152 (56–655) | 136 (52–587) | 0 (0–1833) | 111 (15–614) | 610 (3996–58949) | 3976 (1062–15072) | 3505 (941–13269) |
| South Carolina | 217 (127–469) | 04/15/2020 (04/10/2020–04/18/2020) | 277 (254–777) | 68 (60–173) | 62 (55–158) | 0 (0–0) | 0 (0–0) | 277 (4394–18076) | 2014 (1136–4422) | 1764 (999–3878) |
| South Dakota | 94 (7–378) | 05/10/2020 (04/02/2020–05/08/2020) | 102 (18–501) | 26 (5–128) | 23 (4–114) | 0 (0–0) | 0 (0–54) | 102 (177–14900) | 849 (52–3823) | 748 (47–3362) |
| Tennessee | 231 (136–470) | 04/04/2020 (04/04/2020–04/18/2020) | 282 (267–690) | 71 (69–158) | 65 (63–144) | 0 (0–0) | 0 (0–0) | 282 (4630–17481) | 2104 (1222–4392) | 1853 (1078–3868) |
| Texas | 957 (472–2520) | 04/15/2020 (04/11/2020–04/18/2020) | 1364 (1092–4851) | 308 (242–1025) | 270 (218–933) | 0 (0–0) | 0 (0–0) | 1364 (18370–104777) | 9239 (4451–24648) | 8017 (3875–21418) |
| Utah | 202 (40–753) | 04/24/2020 (04/10/2020–04/25/2020) | 332 (75–1304) | 69 (16–270) | 58 (14–229) | 0 (0–0) | 0 (0–100) | 332 (1805–39658) | 2192 (407–8194) | 1849 (347–6921) |
| Vermont | 40 (33–75) | 04/01/2020 (04/01/2020–04/17/2020) | 54 (48–158) | 14 (13–36) | 12 (12–34) | 0 (0–0) | 0 (0–2) | 54 (995–2825) | 364 (272–702) | 321 (244–617) |
| Virginia | 763 (277–2465) | 04/23/2020 (04/10/2020–04/24/2020) | 1110 (745–4374) | 243 (124–949) | 208 (111–815) | 0 (0–0) | 0 (0–620) | 1110 (12399–114439) | 7849 (2787–25114) | 6706 (2385–21462) |
| Washington | 694 (611–883) | 04/05/2020 (04/05/2020–04/18/2020) | 1043 (1008–1108) | 241 (237–260) | 216 (213–233) | 0 (0–0) | 0 (0–0) | 1043 (22444–35187) | 6515 (5606–8436) | 5692 (4913–7359) |
| West Virginia | 22 (12–58) | 04/14/2020 (04/12/2020–04/18/2020) | 47 (30–170) | 11 (8–39) | 10 (8–37) | 0 (0–0) | 0 (0–0) | 47 (338–2145) | 196 (96–538) | 173 (86–472) |
| Wisconsin | 302 (211–609) | 04/11/2020 (04/11/2020–04/18/2020) | 425 (403–1172) | 108 (99–274) | 95 (90–253) | 0 (0–0) | 0 (0–102) | 425 (7243–22391) | 2766 (1885–5601) | 2432 (1665–4931) |
| Wyoming | 243 (44–948) | 04/30/2020 (04/24/2020–05/01/2020) | 670 (117–3024) | 150 (21–663) | 137 (18–611) | 0 (0–1955) | 106 (0–619) | 670 (803–44921) | 2276 (223–10788) | 1991 (178–9590) |

## Discussion

This study has generated estimates of predicted health service utilisation and deaths due to COVID-19 by day through the end of July for all USA states and EEA countries, assuming that social distancing efforts will continue until deaths reach a very low level. The analysis shows large gaps between need for hospital services and usual capacity, especially for inpatient and ICU beds. A similar or perhaps even greater gap for ventilators is also likely, but detailed state or country data on ventilator capacity are not available to directly estimate that gap. Uncertainty in the time course of the epidemic, its duration, and the peak of utilisation and deaths is large, particularly for when locations are early in the epidemic and where there are few deaths. Given this, it is critical to update these projections as the pandemic progresses and new data are collected. Uncertainty will also be reduced as we gain more knowledge about the epidemic peak and subsequent decline in daily deaths across more than 13 locations. A critical aspect to the size of the peak is when aggressive measures for social distancing are implemented in each state, region, or country and for how long they are maintained. Delays in implementing government-mandated social distancing and relaxing policies will have an important effect on the resource gaps that health systems will be required to manage.

Our estimates of excess demand show that hospital systems have already or will face difficult choices to continue providing high-quality care to their patients in need. This model was first developed for use by the UW Medicine system in Washington state, and the practical experience of that system provides insight into how it has been useful for planning purposes. From the perspective of planning for the UW Medicine system, these projections immediately made apparent the need to rapidly build available capacity. Strategies to do so included suspending elective and non-urgent surgeries and procedures, while supporting surge planning efforts and reconfiguration of medical/surgical and ICU beds across the system. These targets also supported a proactive discussion regarding the potential shift from current standards of care to crisis standards of care, with the goal to do the most good for the greatest number in the setting of limited resources.

There are a variety of options available to deal with the situation, some of which have already been implemented or are being implemented. One option is to reduce non-COVID-19 patient use. In the USA and in many EEA countries, local, state, or national governments have cancelled elective procedures^40–45^ and many, but not all, hospitals have complied. This decision has significant financial implications for USA health systems, however, as elective procedures are a major source of revenue.^46^ Also, aggressive social distancing policies reduce not only the transmission of COVID-19 but will likely have the added benefit of reducing health-care utilisation due to other causes such as injuries.^47^ Reducing non-COVID-19 demand alone will not be sufficient, and strategies to increase capacity are also clearly needed. This includes setting up additional beds by repurposing unused operating rooms, pre- and post-recovery rooms, procedural areas, medical and nursing staff quarters, and hallways.

Currently, one of the largest constraints on effective care may be the lack of ventilators. One supplement to ventilator capacity is using anesthesia machines freed up by deferring or cancelling elective surgeries. Other options go beyond the capacity or control of specific hospitals. The use of mobile military resources has the potential to address some capacity limitations, particularly in the USA given the differently timed epidemics across states. Other innovative strategies will need to be found, including the construction of temporary hospital facilities as has been done in Wuhan,^48^ Washington state,^49^ New York,^50,51^ Italy,^52^ France,^53^ and Spain.^54^

In this study, we have quantified the potential gap in physical resources, but there is an even larger potential gap in human resources (HR). Expanding bed capacity beyond licensed bed capacity may require an even larger increase in the HR to provide care. The average annual bed-day utilisation rate in the US is 66% and ranges from 46%to 92% among EEA countries. Many hospital systems are staffed appropriately at their usual capacity utilisation rate, and expanding even up to, but then potentially well beyond, licensed capacity will require finding substantial additional HR. Strategies include increasing overtime, training operating room and community clinic staff in inpatient care or physician specialties in COVID-19 patient care, rehiring recently separated workers, and the use of volunteers. In academic health systems such as UW Medicine, clinical faculty time can be redirected from research and teaching to clinical care during the COVID-19 surge. A more concerning HR bottleneck identified, given the need for ICU care for COVID-19 patients, is for ICU nurses, for which there are very limited options for increasing capacity. In addition to HR, what should not be overlooked is the increased demand for supplies ranging from personal protective equipment (PPE), medication, and ventilator supplies to basics such as bed linen. Add to these the need to expand other infrastructure required to meet the COVID-19 surge, such as information technology (IT) for electronic medical records. The overall financial cost over a short period of time is likely to be enormous, particularly when juxtaposed against the substantial reductions in revenue for many hospitals due to the cancellation of elective procedures and the broader economic consequences of social distancing mandates.

Our model suggests that the timing of the implementation of social distancing mandates is a critical determinant of peak demand and cumulative deaths. Mobility data derived from cell phone use has provided the basis for evaluating the importance of the different social distancing mandates included in the social distancing covariate. It is important to note the social distancing mandates do not capture all variation in mobility, and that the data in some locations suggests behavioural change prior to the introduction of these mandates. Understanding what drives individual change, e.g. levels of awareness or fear of the pandemic or the private sector implementing remote work policies prior to government mandates, will be important for understanding what may drive the change in behaviour after official social distancing mandates are relaxed, which is now beginning in some European countries and US States.

Based on our experience thus far, we have derived important insights into the epidemic trajectories and health service demand as data have accumulated. These have led to improved forecasts reflecting both new data and method refinements.^55,56^ For this reason, we are continuing to revise the model as new data are available, providing an updated forecast for health service providers, governments, and the public. In some regions that have peaked, such as regions in Italy like Liguria, or New York, the duration of the peak is much longer than in other places such as Madrid. The mixture model we use accommodates this longer peak but it remains unclear why some communities have the prolonged peak and others do not. The prolonged peak leads to substantially increased total mortality. There is also marked variation across locations in how steeply the epidemic curve rises, captured by the alpha parameter in our model. Understanding why some locations have an epidemic like New York and others like Washington State will be important to make robust forecasts in other regions of the world.

Any attempt to forecast the COVID-19 epidemic has many limitations. Only a limited number of locations with generalised epidemics have reached the peak in terms of daily deaths, and only one has currently brought new cases to near zero, namely Wuhan. Many other locations, including all other provinces in China, have so far successfully contained transmission, preventing a general outbreak. Modelling based on one completed epidemic, at least for the first wave, and many incomplete epidemics is intrinsically challenging. The main limitation of our study is that observed epidemic curves for COVID-19 deaths define the likely trajectory. In this study, we do include a covariate meant to capture the timing of social distancing measures and their effect on various measures of population mobility. Our model also relies on the accuracy of reporting of deaths due to COVID-19; reports suggest that in some locations not all deaths may be included in country reported totals.^57,58^ Our models explicitly take into account variation in age-structure, which is a key driver of all-age mortality. But these efforts at quantification do not take into account many other factors that may influence the epidemic trajectory: sex, the prevalence of co-morbidity, population density, individual behavior change not captured by mobility metrics, and a host of other individual factors that may potentially influence the immune response. We also have not explicitly incorporated the effect of reduced quality of care due to stressed and overloaded health systems beyond what is captured in the data. For example, the higher mortality rate in Italy may in part be due to policies around restricting invasive ventilation in the elderly. The model ensemble used does suggest that locations with faster increases in the death rate are likely to have greater peak caseload and cumulative deaths, but our uncertainty intervals are appropriately large. Finally, it is critical to note that we restrict our projections to the first wave of the pandemic under a scenario of continued implementation of social distancing mandates and do not yet incorporate the possibility of a resurgence or subsequent waves. This is an essential area for future work.

## Conclusion

COVID-19 is an extraordinary challenge to health and the health-care system. In this study, we forecast a large excess of demand for hospital bed-days and ICU bed-days and our estimate of 1,584,737 (95% UI 1,050,954–3,082,999) deaths in the USA and EEA from the first wave of pandemic is an alarming number. This number could be substantially higher if excess demand for health system resources is not addressed and if social distancing policies are not continued, vigorously implemented, and enforced. This planning model will hopefully provide an up-to-date tool for improved hospital resource allocation.

## Data Availability

Additional information on the determination of hospital resource utilisation and capacity is provided in Appendix A; details on curve fitting methods, quantification of uncertainty, and a full specification of the statistical model are available in Appendix B. This study complies with the Guidelines for Accurate and Transparent Health Estimates Reporting (GATHER) statement.

https://covid19.healthdata.org/

